# Marburg Virus Disease outbreaks, mathematical models, and disease parameters: a Systematic Review

**DOI:** 10.1101/2023.07.10.23292424

**Authors:** Gina Cuomo-Dannenburg, Kelly McCain, Ruth McCabe, H. Juliette T. Unwin, Patrick Doohan, Rebecca K. Nash, Joseph T. Hicks, Kelly Charniga, Cyril Geismar, Ben Lambert, Dariya Nikitin, Janetta Skarp, Jack Wardle, Pathogen Epidemiology Review Group, Mara Kont, Sangeeta Bhatia, Natsuko Imai, Sabine van Elsland, Anne Cori, Christian Morgenstern

## Abstract

**Background:** Recent Marburg virus disease (MVD) outbreaks in Equatorial Guinea and Tanzania highlighted the importance of better understanding this highly lethal infectious pathogen. Past epidemics of Ebola, COVID-19, and other pathogens have re-emphasised the usefulness of mathematical models in guiding public health responses during outbreaks.

**Methods:** We conducted a systematic review, registered with PROSPERO (CRD42023393345) and reported according to PRISMA guidelines, of peer-reviewed papers reporting historical outbreaks, modelling studies and epidemiological parameters focused on MVD, including contextual information. We searched PubMed and Web of Science until 31st March 2023. Two reviewers evaluated all titles and abstracts, with consensus-based decision-making. To ensure agreement, 31% (13/42) of studies were double-extracted and a custom-designed quality assessment questionnaire was used to assess the risk of bias.

**Findings:** We present detailed outbreak, model and parameter information on 970 reported cases and 818 deaths from MVD until 31 March 2023. Analysis of historical outbreaks and sero-prevalence estimates suggests the possibility of undetected MVD outbreaks, asymptomatic transmission and/or cross-reactivity with other pathogens. Only one study presented a mathematical model of MVD transmission. We estimate an unadjusted, pooled total random effect case fatality ratio for MVD of 61.9% (95% CI: 38.8-80.6%, *I*^2^=93%). We identify key epidemiological parameters relating to transmission and natural history for which there are few estimates.

**Interpretation:** This review provides a comprehensive overview of the epidemiology of MVD, identifying key knowledge gaps about this pathogen. The extensive collection of knowledge gathered here will be crucial in developing mathematical models for use in the early stages of future outbreaks of MVD. All data are published alongside this article with functionality to easily update the database as new data become available.

**Funding:** MRC Centre for Global Infectious Disease Analysis

**Research in Context:**

- **Evidence before this study** We searched Web of Science and PubMed up to 31 March 2023 using the search terms Marburg virus, epidemiology, outbreaks, models, transmissibility, severity, delays, risk factors, mutation rates and seroprevalence. We found five systematic reviews, all of which considered MVD alongside Ebola virus disease (EVD). One modelling study of Marburg virus disease (MVD) focused on animals, and not on computational models to understand past or project future disease transmission. One systematic review collated risk factors for transmission based on four MVD studies, but did not report attack rates due to missing underlying MVD estimates; another systematic review pooled estimates of MVD case fatality ratios (CFR): 53.8% (95% CI: 26.5–80.0%) and seroprevalence: 1.2% (95% CI: 0.5–2.0%). No systematic review covered transmission models of MVD, and the impact of public health and social measures is unknown.
- **Added value of this study** We provide a comprehensive summary of the available, peer-reviewed literature of historical outbreaks, transmission models and parameters for MVD. Meta-analysis of existing estimates of CFRs, and our original estimates based on historical outbreak information, illustrate the severity of MVD with our pooled random effect estimated CFR of 61.9% (95% CI: 38.8-80.6%, *I*_2_=93%). We demonstrate the sparsity of evidence on MVD transmission and disease dynamics, particularly on transmissibility and natural history, which are key input parameters for computational models supporting outbreak response. Our work highlights key areas where further disease characterization is necessary.
- **Implications of all the available evidence** Previous outbreaks of infectious pathogens emphasized the usefulness of computational modelling in assessing epidemic trajectories and the impact of mitigation strategies. Our study provides necessary information for using mathematical models in future outbreaks of MVD, identifies uncertainties and knowledge gaps in MVD transmission and natural history, and highlights the severity of MVD.

## 1 Introduction

Infectious disease outbreaks pose a substantial threat to health and well-being globally [1, 2, 3]. Since the emergence of SARS-CoV-2 at the end of 2019, there have been several other outbreaks of emerging or reemerging pathogens, including mpox ([4]), novel hepatitis in children ([5]), Ebola virus disease (EVD) ([6]), and Marburg virus disease (MVD) ([7, 8]). These examples demonstrate that the world remains highly vulnerable to infectious disease outbreaks and underscores the importance of developing a better understanding of high-threat pathogens.

In 2018, the World Health Organization (WHO) published a list of nine known pathogens for research and development (R&D) prioritisation, due to their epidemic and pandemic potential and the absence of licensed vaccines or therapeutics [9]. Among these is Marburg virus (MV), a highly-lethal infectious *Filoviridae* single-stranded RNA virus, first described in Germany and Serbia (formerly Yugoslavia) in 1967. Subsequent outbreaks of MVD have primarily occurred in sub-Saharan Africa, including recent outbreaks in Equatorial Guinea and Tanzania in 2023 [7, 8].

The host of MV is the fruit bat (*Rousettus aegyptiacus*), with human transmission occurring via direct contact with an infected animal host or an infected human [10, 11]. Phylogenetic analyses have confirmed multiple spillovers from bats to humans [12], but the first known human outbreak was associated with African green monkeys (*Cercopithecus aethiops*) [13]. Clinical symptoms include, but are not limited to, fever, severe headaches and malaise, which can progressively develop into severe hemorrhagic fever, including spontaneous bleeding from one or more orifices [13], with a high risk of serious illness upon infection [12]. The European Centre for Disease Prevention and Control MVD factsheet provides a more comprehensive overview [14].

Mathematical models of disease transmission and control are a key tool that can be deployed in response to infectious disease outbreaks and are used to guide policy, for example by projecting plausible epidemic trajectories and expected healthcare demand and assessing the potential impact of interventions [15, 16]. Epidemiological parameters are key inputs to such models, for example governing the rates at which individuals move through disease states. However, gathering information on model structures and appropriate parameter values can be time-consuming and may impede real-time modelling.

To address these issues, we have set out to systematically review the literature relevant to rapid design of dynamic transmission models for priority pathogens. We aim to collate available information on outbreaks, modelling studies, and epidemiological parameters related to transmissibility, severity, delays, risk factors, mutation rates and seroprevalence for each of the nine aforementioned priority pathogens [9]. Our work will highlight knowledge gaps and provide a key resource for modelling future outbreaks of these or similar (known or unknown) pathogens. This paper is the first in a series from this project, presenting results of our systematic review of MVD.

## 2 Methods

PRISMA checklists for this review have been included in Tables S5 and S6.

### Search strategy and study selection

We searched for published mathematical transmission models and articles reporting on MVD transmission, evolution, natural history, severity, seroprevalence and size of previous outbreaks, published prior to 31 March 2023 (see Supplement A.1 for search strategy). Table S1 presents all inclusion and exclusion criteria. In *Covidence* [17], two independent reviewers screened titles and abstracts then full texts to assess eligibility for data extraction. Disagreements were resolved by consensus between reviewers.

### Data extraction

Thirteen reviewers extracted data on article information (publication details, risk of bias), estimated parameters (value, uncertainty range, distribution, context, risk factors), outbreaks (dates, location, case and death numbers) and models (model type and structure, interventions modelled, transmission routes, assumptions) from the included studies into a Microsoft Access database (Version 2305), with one reviewer per paper. Risk of bias was assessed using a seven-question form addressing methodology, assumptions and data. For a randomly selected 30% (13/43) of papers, extraction was performed by two independent reviewers. Consensus on discordant results was established before single reviewer data extraction commenced. More details are provided in Supplements S1, S3.

We only collated information from outbreaks that were reported to be complete.

We extracted parameter values, units, uncertainty intervals (capturing the precision of estimates), and ranges (capturing heterogeneity in estimates across different population groups, time or space) for all parameters except risk factors. Study context was also recorded, when reported. We extracted risk factors investigated in the studies and whether they were statistically significant and/or adjusted. We chose not to extract odds ratio estimates because varying stratifications and reference groups complicates comparison across studies.

Information extracted about previous outbreaks, namely cases and deaths, was further used to generate estimates of the case fatality ratio (CFR).

Full details on data extraction, including descriptions of variables and predefined options for categorical variables, can be found in the Supplement S2 S3 S4C.

### R package

We designed an R package, *epireview*, where all curated data on epidemiological parameters, models and outbreaks are publicly available [18]. A dedicated vignette explains how independent contributors may add information to the package, so that it provides a live view of the latest knowledge on MVD. More details can be found in Supplement C.

### Data analysis

We use descriptive tables and figures to present the collated data. Unless otherwise specified, uncertainty intervals in tables and figures (e.g. 95% confidence (CI) or credible intervals (CrI)) were extracted from the papers or computed from reported central estimates and standard errors (A.3).

In the following, an “unadjusted CFR estimate” refers to an estimate where raw deaths are divided by raw cases, with no weighting or controlling for other variables or cases with unknown outcome.

We conducted two meta-analyses for the case fatality ratio (CFR), using 1) CFR estimates extracted from the studies, and 2) unadjusted CFRs that we computed from extracted outbreak data. Comparison between the two sets of CFR estimates enabled to assess any bias due to outbreaks for which there was no or multiple reported CFR estimates in the literature. For this analysis, we defined an ‘outbreak’ as one or more cases identified in the same country and within the same date ranges. This included single cases, often related to zoonotic spillover or importation events, and large outbreaks. We ensured that each case was counted only once: if multiple studies reported the same outbreak, we chose the study covering the longest time period. We estimated exact 95% binomial confidence intervals on individual outbreak estimates.

Meta-analyses were performed using the `meta` R package [19] providing a *total common effect* and a *total random effect* pooled CFR estimate with 95% CI and statistics on heterogeneity in CFR across studies. Further details and references are provided in Supplement A.4.

Overall quality assessment scores were calculated as a mean of the responses to the seven questions, excluding non-applicable questions (that is, if the quality assessment question was not applicable to a study, it did not contribute the the quality assessment score). A local polynomial regression fit using the *R* function *loess* was used to analyse trends in quality assessment scores by publication year.

Analyses were conducted using *R* (version 4.2.2).

## 3 Results

### Study Selection

The search returned 4410 studies (2305 from PubMed and 2105 from Web of Science) from which we removed 1256 duplicates. Of the remaining 3154 studies for which we screened abstracts, 221 were kept for full-text review. Studies were further excluded for various reasons, including not reporting any parameter or original parameter estimates, not being peer-reviewed, being duplicated, and being in a non-English language. 42 studies were included for data extraction. The PRISMA flowchart further describes the study selection (Figure

### Historical Outbreaks

We collated evidence from 13 studies reporting 23 observed MVD outbreaks. Based on timings and locations reported in the studies, we identified seven distinct outbreaks (Table 1). This included the first identified outbreak in Marburg, Germany, and the former Federal People’s Republic of Yugoslavia from which MVD was identified and named; an outbreak in the Democratic Republic of the Congo (DRC) from 1998 - 2000; a series of cases from Johannesburg, South Africa in early 1975 (linked to prior travel to Zimbabwe); three outbreaks in Uganda; and an outbreak in Angola in 2004 - 2005. In addition, we noted the reporting of individual MVD cases in Kenya in 1980 and 1987 (likely caused by animal exposure); in the Russian Federation in 1988 and 1990 (both linked to a laboratory worker in a research facility); and in the Netherlands and the United States of America in 2008, both linked to the 2007 Ugandan outbreak. At the time of the literature search, there were no peer-reviewed studies on the 2023 MVD outbreaks in Equatorial Guinea and Tanzania.

**Table 1:**
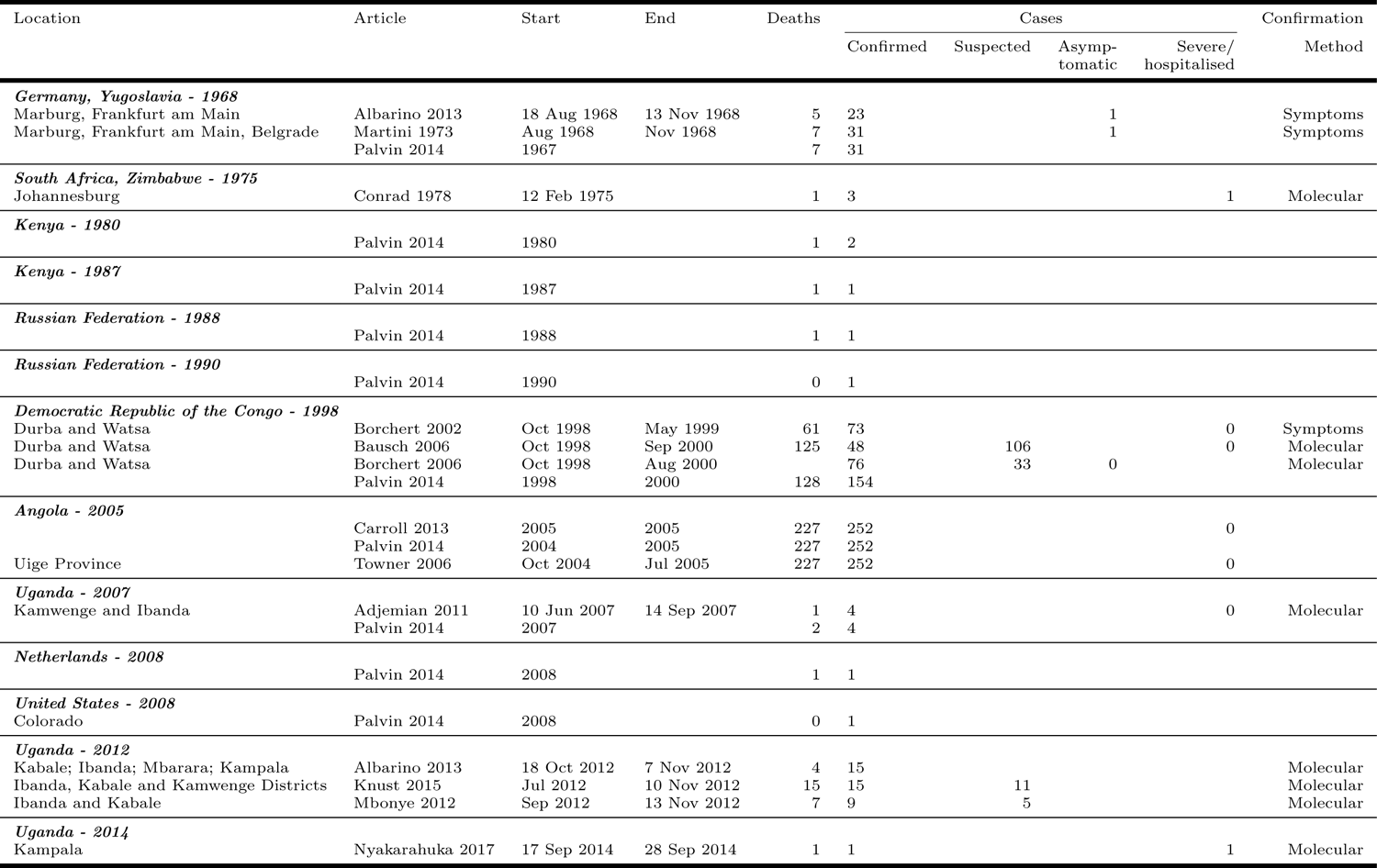
Overview of MVD outbreaks i.e. location, timing, and size, as reported in the studies included in this review. We report in bold the country and outbreak year, the location refers to the place of the actual outbreak in the country if known. Blank cells correspond to information which we were unable to find in or extract from the literature.

### Mathematical Models

Ajelli et al, 2012 was the only MVD transmission modelling study. The authors used a stochastic, individual-based, SEIR model to examine the impact of behaviour change interventions on MVD cases and deaths [20]. Transmission in the model occurred via direct, non-sexual human contact, assuming homogeneous mixing; transmission rates were heterogeneous over time, with temporal changes in viral load and hence transmissibility; susceptibility was assumed to be age-dependent, and the latent and incubation periods were assumed to coincide [20]. The potential impact of quarantine was simulated, though was not explicitly based on real-world data. As detailed below, the authors provided estimates of generation time and basic reproduction number [20].

### Epidemiological Parameters

We extracted 71 parameter estimates: see overview in Figure S1and parameter definitions and details of the extraction process in the accompanying R package *epireview* 18.

Seroprevalence estimates were the most frequently reported in the literature, followed by delays and severity. Two studies reported on transmission parameters (e.g., attack rates and reproduction numbers), and four provided estimates of evolutionary mutation rates. We also extracted reported risk factors for different outcomes, namely infection, severe disease, seropositivity, recovery, and death.

### Transmission

Two studies reported reproduction number estimates [21, 20]. Ajelli et al. used a mathematical model (see Section 3), to estimate the basic reproduction number, *R*_0_, for the 2005 Angola outbreak. They found that *R*_0_ = 1.59 (95% CI: 1.53–1.66), suggesting that in the absence of mitigation efforts, the virus would be expected to propagate in a similar population [20]. They also provided the only estimate of doubling time, at 12.4 days (95% CI: 11.3–13.6 days)[20].

Borchert et al. estimated the effective reproduction number, *R_e_*, based on secondary attack rates derived from seroprevalence in contacts of confirmed cases in DRC in 2002 [21]. This study also provided the only estimate of attack rate, at 21% (Figure 3).

**Figure 1:**
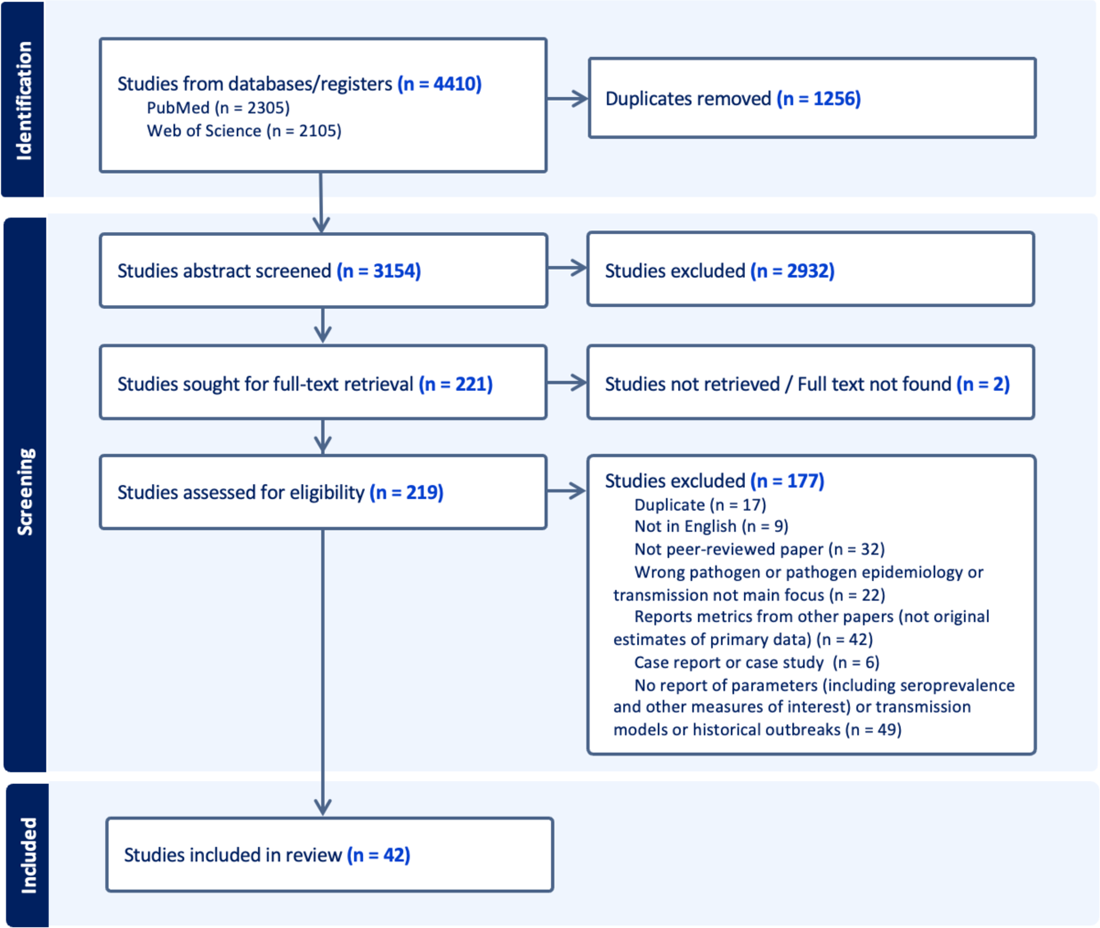
Study selection according to PRIMSA guidelines and criteria as described in Table S1

**Figure 2:**
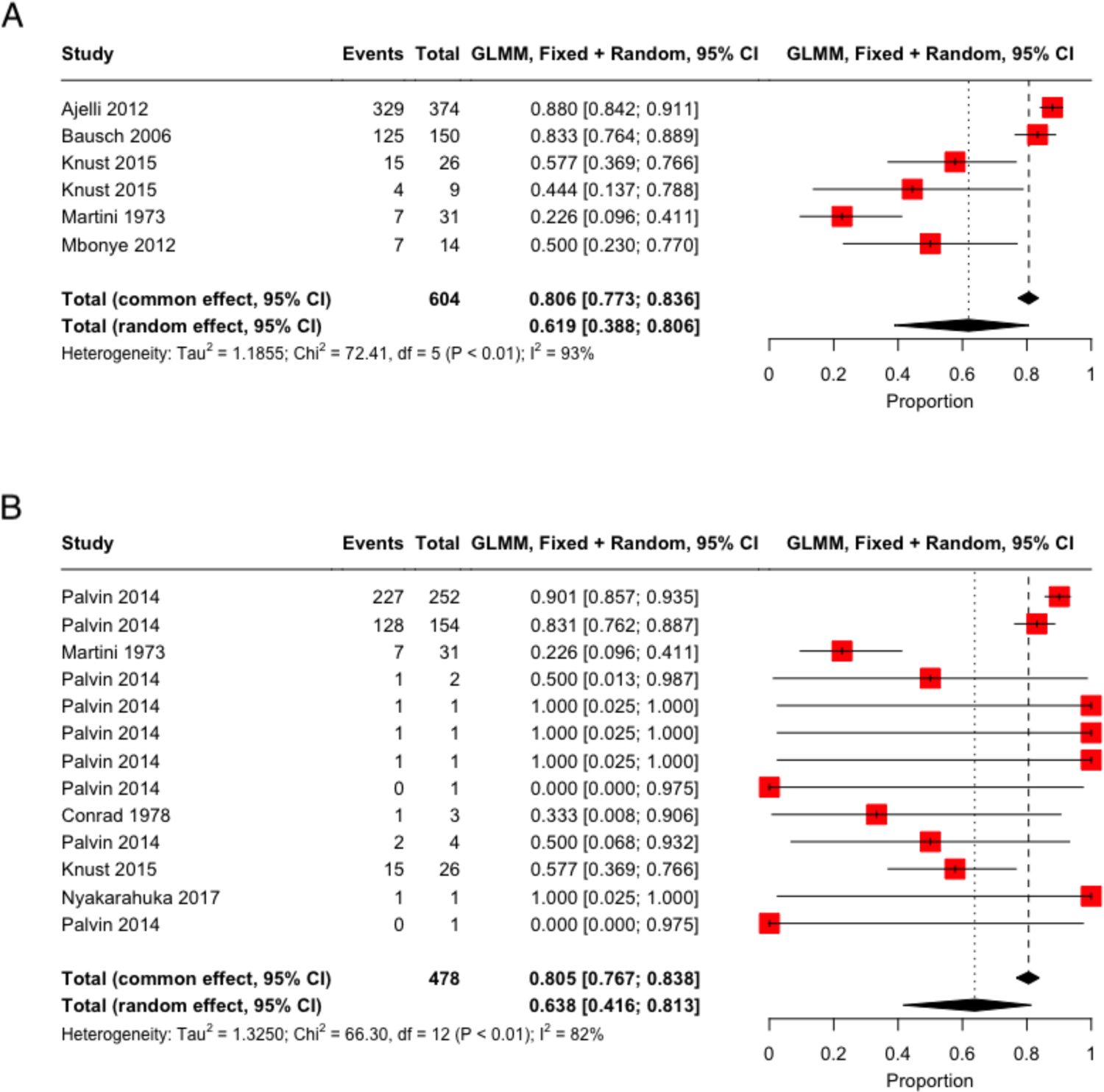
Case Fatality Ratio (CFR) meta-analyses, using logit-transformed proportions and a generalized linear mixed-effects model (GLMM) (full details in SI A.4). The forest plot displays studies included in each meta-analyis: the red squares indicate study weight, and for each study, a 95% binomial confidence interval is provided. To summarize, we display as black diamonds the *total common effects*, where all data are effectively pooled and assumed to come from a single data-generating process with one common CFR and *total random effect* estimates, which allow the CFR to vary by study and accordingly give different weights to each study when determining an overall estimate [19]. (A) CFR estimates reported in the included studies. (B) CFR estimated from extracted outbreak data, including only one observation per outbreak using the study with the longest duration of the outbreak reported ensuring no case is double counted.

**Figure 3:**
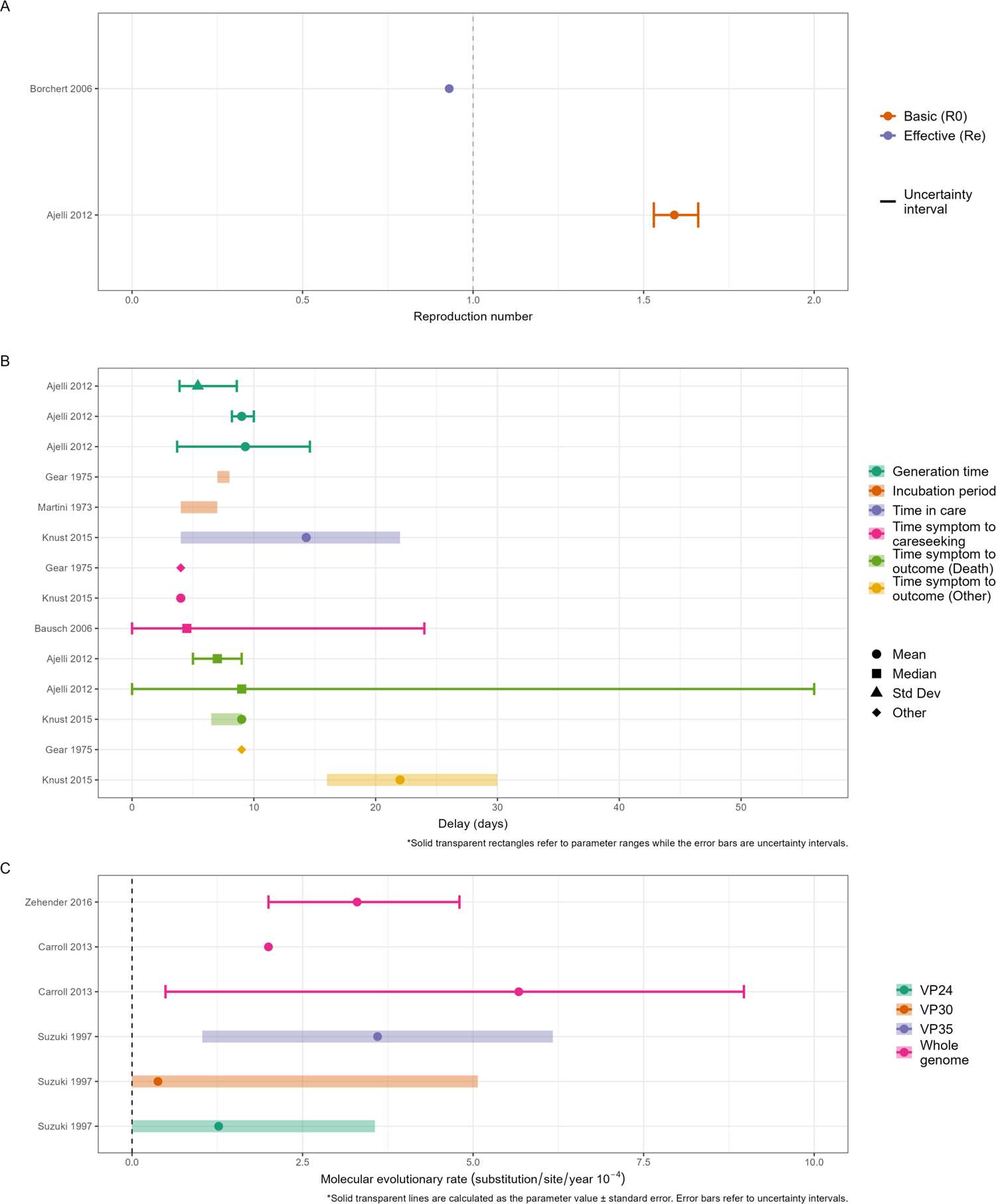
Overview of the reproduction numbers, delays and evolutionary rate estimates from the included studies of MVD. Solid lines represent uncertainty intervals and ribbons indicate a parameter range (e.g. across different populations or over time). (A) Estimates of the reproduction number. The blue and red points correspond to estimates of the effective reproduction number (*R_e_*) and basic reproduction number (*R*_0_) respectively, with associated uncertainty shown by the solid lines where available. The dashed vertical line presents the threshold for epidemic growth. (B) Delay parameters, stratified into five categories: Generation Time, Incubation Period, Time in Care, Time from Symptom to Careseeking and Time from Symptom to Outcome as indicated by different colours. (C) Evolutionary rates. Colours indicate different genome types; points represent central estimates. Solid lines represent an uncertainty interval associated with the point estimate while ribbons indicate a parameter value +/- standard error with a minimum set to zero.

### Severity

Six CFR estimates were reported, corresponding to the outbreaks in Angola in 2005 [20], DRC in 1999 [12], the original 1968 outbreak in Germany and Yugoslavia [22] and three estimates from the 2012 Uganda outbreak [23] [24] (Figure S2A). Pooling these estimates gave a total common effect CFR of 80.6% (95% CI: 77.3-83.6%, *I*^2^=93%) and a total random effect CFR of 61.9% (95% CI: 38.8-80.6%, *I*^2^=93%).

We additionally estimated an unadjusted, pooled CFR using the extracted historical outbreak data (Figure S2B), combining data from 467 confirmed cases and 11 suspected cases across 13 distinct outbreaks with 385 reported deaths. The pooled common effect CFR estimate from the extracted outbreak data was 80.5% (95% CI: 76.7-83.8%, *I*^2^=82%) and the pooled random effect CFR 63.8% (95% CI: 41.6-81.3%, *I*^2^=82%), both highly consistent with the previous estimates based on CFR parameters reported in the literature.

### Delays

We collated estimates of the generation time, incubation period, time in care, and time from symptom onset to careseeking, death or other outcomes as summarised in Figure 3 and Table 2. The two generation time estimates were based on viral load data from non-human primates under two distinct assumptions, namely that infectiousness is directly proportional to viral load, and alternatively assuming that probability of death is directly proportional to viral load [20, 25]. This study also estimated the time from symptom onset to death using additional assumptions about these relationships [20]. The sole estimate of time in care was a median of 14.3 days (range 4 - 22 days) that 6 survivors of the 2012 Uganda outbreak spent in care, with a median duration in isolation of 22 days (16 - 30 days) [23]. The two incubation period estimates came from studies from the 1970s only reporting ranges with little overlap [26, 22] (Figure 3). Central estimates of time from symptom onset to careseeking across the 1975 South Africa, 1998 DRC, and 2012 Uganda outbreaks were consistently under 5 days, although Bausch et al. showed a large range of delays from symptoms to seeking medical care for the 1998 DRC outbreak.

**Table 2:** Overview of the MVD delay parameter estimates extracted from the included studies. These are stratified into five categories: Generation Time, Incubation Period, Time in Care, Time from Symptom to Careseeking and Time from Symptom to Outcome. Estimates and associated uncertainty are provided, along with information regarding the population group corresponding to the estimate and the timing and location of the outbreak. ‘Other’ refers to a range of different values which are specified in the underlying papers.

| Article | Country | Survey year | Delay (days) | Statistic | Uncertainty (days) | Uncertainty type | Population Group | Timing of survey | Outcome |
| --- | --- | --- | --- | --- | --- | --- | --- | --- | --- |
| **Generation Time** |  |  |  |  |  |  |  |  |  |
| Ajelli 2012 # | Angola | Mar-Nov 2005 | 9.3 | Mean | 3.7 - 14.6 | CI95% | General population | Mid outbreak |  |
| Ajelli 2012 | Angola | Mar-Nov 2005 | 5.4 | Standard Deviation | 3.9 - 8.6 | CI95% | General population | Mid outbreak |  |
| Ajelli 2012 # | Angola | Mar-Nov 2005 | 9 | Mean | 8.2 - 10 | CI95% | General population | Mid outbreak |  |
| **Incubation Period** |  |  |  |  |  |  |  |  |  |
| Martini 1973 | Germany, Yugoslavia | Aug-Nov 1968 |  |  | 4 - 7 | Range |  |  |  |
| Gear 1975 | South Africa | Feb 1975 |  |  | 7 - 8 | Range | Healthcare workers | Mid outbreak |  |
| **Time In Care** |  |  |  |  |  |  |  |  |  |
| Knust 2015 | Uganda | 2012 | 14.3 | Mean | 4 - 22 | Range | General population | Post outbreak | Other |
| **Time From Symptom To Careseeking** |  |  |  |  |  |  |  |  |  |
| Bausch 2006 | DRC* | 1999-2000 | 4.5 | Median | 0 - 24 | Range | Other | Mid outbreak |  |
| Gear 1975 | South Africa | Feb 1975 | 4 | Other |  |  | Other | Start outbreak |  |
| Knust 2015 | Uganda | 2012 | 4 | Mean |  |  | General population |  |  |
| **Time From Symptom To Outcome** |  |  |  |  |  |  |  |  |  |
| Ajelli 2012 | Angola | Mar-Nov 2005 | 7 | Median | 5 - 9 | Range | General population | Mid outbreak | Death |
| Ajelli 2012 | Angola | Mar-Nov 2005 | 9 | Median | 0 - 56 | Range | General population | Mid outbreak | Death |
| Gear 1975 | South Africa | Feb 1975 | 9 |  |  |  | Other | Start outbreak | Other |
| Knust 2015 | Uganda | 2012 | 9 | Mean | 6.5 - 9 | Range | General population | Post outbreak | Death |
| Knust 2015 | Uganda | 2012 | 22 | Mean | 16 - 30 | Range | General population |  | Other |

\*DRC: Democratic Republic of the Congo

**Table 3:** Aggregated information on risk factors associated with MVD infection and seropositivity. Risk factors were mapped onto our risk factor classification (see Supplement) by interpreting the authors’ descriptions. Adjusted refers to whether estimates were adjusted (i.e. from a multivariate analysis) or not (i.e. from a univariate analysis), with unknown showing that this information is not clearly stated in the original study. Statistical significance was determined according to the original authors’ statistical approaches when specified, or using a p-value of 0.05 otherwise. The numbers in the significant and not significant columns represent the total sample size included in the analyses for this risk factor and outcome category.

| Risk factor | Adjusted | Sample size (Significant) | Sample size (Not significant) |
| --- | --- | --- | --- |
| ***Infection*** |  |  |  |
| Contact with animal | Unknown | Unknown |  |
| Gathering | Unknown | 128 |  |
| Household contact | Unknown | 102 |  |
| Occupation - Funeral and burial services | Unknown | 102 |  |
| Other | Unknown | 102 | 26 |
| Sex | Unknown |  | 26 |
| ***Seropositivity*** |  |  |  |
| Contact with animal | Adjusted |  | 912 |
| Contact with animal | Unknown |  | 300 |
| Gathering | Unknown |  | 300 |
| Hospitalisation | Adjusted | 915 |  |
| Household contact | Adjusted |  | 912 |
| Occupation - Funeral and burial services | Adjusted |  | 912 |

#### Risk Factors

15 risk factors for MVD infection and seropositivity were extracted from 4 studies and are presented in Table S1

Having had contact with confirmed MVD cases, including through working in funeral and burial services, was a statistically significant risk factor for infection. The ‘other’ classification encompassed a wide range of factors, such as prevalence of infection in the host reservoir, subsistence activities and previous invasive medical treatment, and as such are not directly comparable, although some constituted statistically significant risk factors [10, 27, 23]. Sex was not significantly associated with MVD infection [23].

Although similar risk factors were explored to assess impact on seropositivity, the only significant risk identified for this outcome was known hospitalisation with MVD.

### Molecular evolutionary rates

Three studies reported molecular evolutionary rates of MV, two estimated using whole genome sequencing [28, 29] and one based on individual genes [30]. The three evolutionary rate estimates from whole genomes are largely consistent with one another, whilst those based on individual genes tended to be lower (Figure 3C).

#### Seroprevalence

Twenty-one studies contained seroprevalence estimates across a 38-year period from 1980 - 2018 in 15 predominantely Sub-Saharan African countries[31, 27, 21, 48, 32, 37, 38, 39, 40, 42, 43] (Table 4). Presence of antibodies were assessed using a range of assays: Indirect Fluorescent Antibody assay (IFA) (6 studies [49, 44, 27, 32 38, 36); Hemagglutination Inhibition Assay (HAI/HI) (1 study 43); Immunoglobulin G (IgG) (7 studies 31, 48, 47, 21, 33, 42, 50); Immunoglobulin M (IgM) (2 studies 39 40); the remaining studies did not specify this information (3 studies [35, 22, 41]). IgG and IgM were used for all recent studies (from 1995 onwards), highlighting recent developments in serology and the retiring of assays testing for IFA and HAI/HI.

**Table 4:**
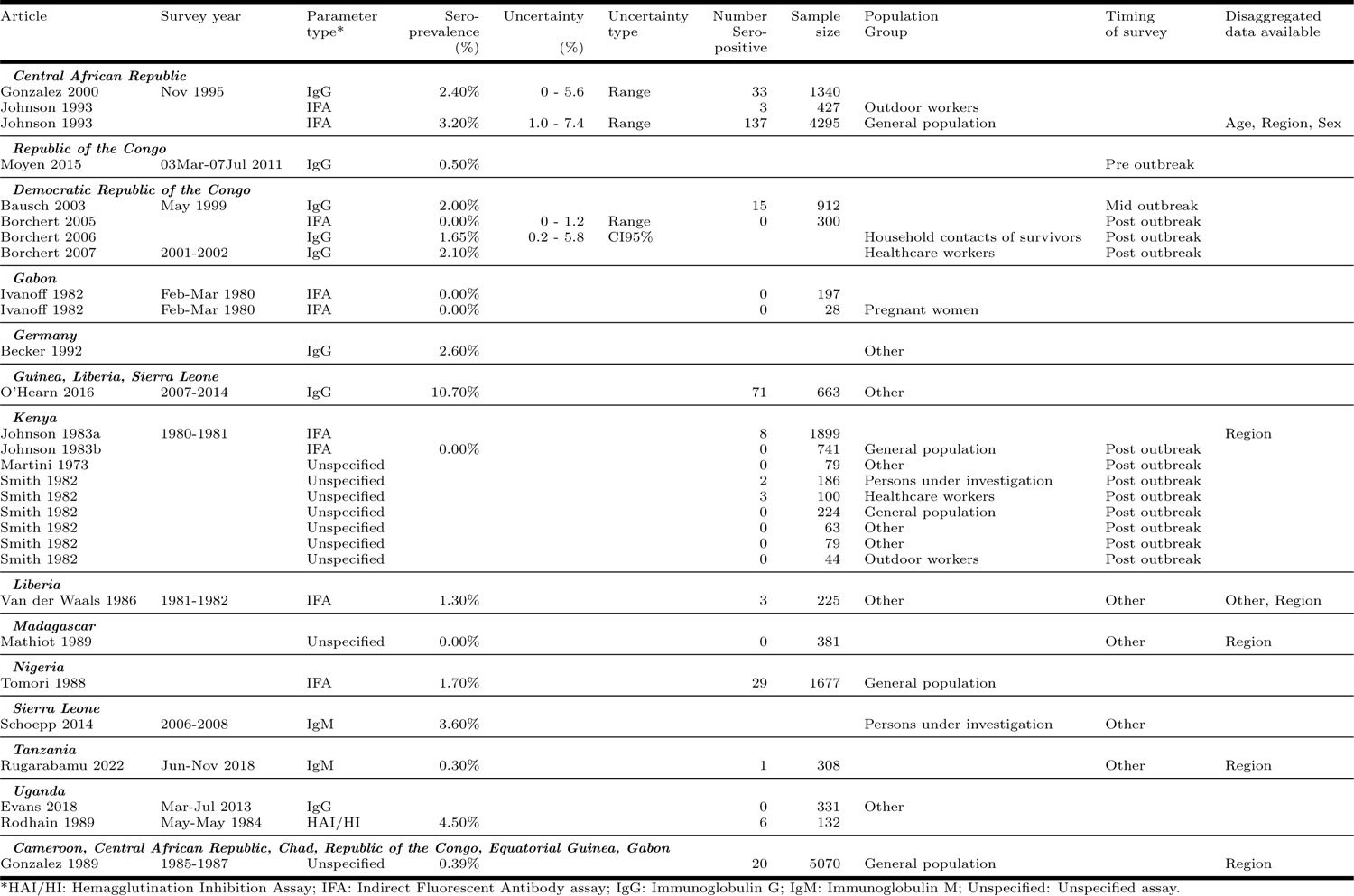
Overview of seroprevalence estimates for MVD as reported in the included studies. Estimates were primarily reported as percentages, as shown here. Associated uncertainty and sample sizes are provided where these were reported. Where available, additional information regarding the location and timing of the estimates, the antibody being tested for, the target population, the timing in relation to any ongoing outbreak and the availability of disaggregated data is also summarised.

The studies included in this review demonstrated low levels of antibodies in surveyed populations, with approximately one third of studies reporting a seroprevalence of 0% [27, 32, 46, 22, 35, 37, 42]. Among studies with estimates above zero, seroprevalence ranged from 0.5% in the Republic of the Congo in 2011 [50], to 2.1% in healthcare workers in DRC in 2001 - 2002 [48], to 4.5% in Uganda in 1984 [43]. Overall, the evidence gathered here indicates high suspectibility to MVD among populations in the surveyed regions, including Tanzania, where one of the subsequent 2023 MVD outbreaks occurred [40]. However, these seroprevalence estimates must be interpreted in the context of the very small sample sizes of most studies.

### Quality assessment

The results of the quality assessment are summarised in (Figure S3A). The number of non-applicable answers are driven by more descriptive studies, such as seroprevalence studies, which did not use a model or statistical analysis. Papers on transmission parameters had on average the highest quality assessment scores (reproduction number paper score = 0.800, other transmission parameters papers score = 0.87, we note the small number of papers in this category) and papers on seroprevalence the lowest score of 0.48. Scores improved over time (Figure S3B) which may also explain the difference in quality assessment score between types of parameters, as seroprevalence papers tended to be published much earlier than other study types.

## 4 Discussion

This systematic review presents a comprehensive set of mathematical models, outbreaks, and epidemiological parameters of MVD. This is the first of a series of systematic reviews covering the 9 WHO priority pathogens listed in 2018.

Historic outbreaks and case reports in the peer-reviewed literature for MVD were rare and small in size, relative to many other pathogens, including other viral hemorrhagic fevers such as Ebola virus disease (EVD), with only 7 notable outbreaks reported (Table 1). Only two outbreaks had over 100 confirmed cases (DRC 1998: 154 cases, Angola 2005: 254 cases), with the remainder reporting 31 cases or fewer. For most parameters, we were only able to obtain a small number of estimates, a substantial number of which were only reported as point estimates with no uncertainty. Seroprevalence of MVD was the metric most widely reported across a large number of locations in Sub-Saharan Africa (Table 4) and indicates that seroprevalance is generally low. However, serosurveys suggest that some past MVD outbreaks may have gone undetected. Reported seroprevalence in the Central African Republic (CAR) is relatively high (3.2%, range among subgroups: 1.0-7.4%) despite having no recorded MVD outbreak, although these results may stem from cross-reactivity or low assay specificity. Seroprevalence estimates of MVD and EVD are often reported together, with estimates for MVD consistently lower than for EVD, e.g. [50, 31, 32, 40].

A basic reproduction number of 1.59 (95% CI: 1.53-1.66) was estimated for the largest known outbreak to date in Angola [20]. However, Borchert et al. [21] estimated an effective reproduction number, *R_e_*, of 0.93 for the 1998 DRC outbreak after the introduction of public health and social measures (PHSM), suggesting that such interventions can effectively mitigate MVD transmission.

The pooled CFR estimates for MVD provide several key insights. The pooled random effects CFR of 61.9% (95% CI: 38.8-80.6%, *I*^2^=93%) highlights the heterogeneity in CFR across outbreaks. In comparison, the pooled common effect CFR of 80.6% (95% CI: 77.3-83.6%, *I*^2^=93%) is skewed towards the two large outbreaks in Angola and Uganda, which had very high CFRs, and presents a possibly misleadingly narrow uncertainty interval but highlights that MVD outbreaks with higher transmissibility may also be associated with higher severity. The results from the meta-analyses of reported CFR parameters and computed, unadjusted CFR from outbreak data are consistent, and our estimates are in line with a previous systematic review [51]. All CFR estimates, irrespective of the method, are extremely high, implying very high costs of human life in the endemic countries, so far all located in Sub-Saharan Africa. Low seroprevalence estimates in these regions, combined with high fatality and a basic reproduction number above one, clearly demonstrate the pandemic potential of MVD.

The gaps in knowledge of MVD are substantial. Although we found some epidemiological estimates, a number of them are from the previous century and based on poor-quality data; for example, most estimates of the CFR for MVD reported in the literature were unadjusted estimates. Crucial model inputs, such as the generation time, were estimated from primate studies and would benefit from confirmation from human outbreak data. Recent outbreaks of MVD in Equatorial Guinea and Tanzania were controlled through basic measures such as *Infection Prevention and Control* and *Risk Communication and Community Engagement* [52]. WHO declared the end of the Equatorial Guinea outbreak on 8 June 2023 [53] (17 laboratory-confirmed cases, 12 deaths, and a further 23 probable cases, all of whom died) and the Ministry of Health of the United Republic of Tanzania confirmed the end of the first MVD outbreak in Tanzania on 2 June 2023 [54] (8 laboratory-confirmed cases, 1 probable case and 6 deaths). These are severe and traumatic events for the communities impacted but also opportunities to gather higher-quality data. In particular, careful collection of patient information, documentation of disease progression and regular follow-ups post-infection would enable the research community to better characterise epidemiological delays and risk factors for infection and death.

The collection of parameters presented here, synthesising peer-reviewed information up to March 2023, will enable researchers to construct and parameterise simple epidemiological models for MVD. Our accompanying R package *epireview* [18] will facilitate this process and ensure that information from studies beyond March 2023 can be added to the package, thereby offering a continuously updated repository of parameter estimates.

The importance of this work is underlined by the scarcity of published MVD mathematical models, which contrasts with the abudance of published models describing EVD [55].

Improved knowledge of parameters will enable more modelling analyses to explore the potential impact of interventions such as PHSM, as has been done for EVD [56]. Although there is no vaccine approved for MVD, phase 1 clinical trials have shown promising results [vaccination strategies, as they did for EVD [58]. Mathematical models could support the design of This review was challenging as it contained a wide variety of studies and parameters for which we could not find a unique pre-existing, validated quality assessment tool. We therefore constructed a scoring system to assess the validity of the methods, assumptions and data, tailored specifically to the broad range of information we were collating. We observed an improvement in paper quality over time, which we attribute to increasing transparency in models, assumptions and data (including publication of data and code), which enables reproducibility of research.

Our findings are limited by our restriction to peer-reviewed articles in English; extending this work to include non-English language articles and non-peer reviewed work is an interesting avenue, but would be challenging.

Although we excluded systematic reviews from our systematic search, we used them (e.g. Nyakarahuka et al [51]) to validate our extracted data.

Lowering the hurdles for mathematical epidemic model design is important to enable timely generation of evidence that can support epidemic response to future outbreaks. Here, we provide a comprehensive summary of published mathematical models, outbreaks, and epidemiological parameters of MVD. Our work summarises existing information on MVD dynamics and highlights key knowledge gaps which would benefit from further elucidation. We publish the database of extracted models, parameters and outbreaks, thus enabling future additions as more information becomes available from future studies. Information is synthesised in the R package epireview [18], which also includes functionalities to visualise the latest information, thereby providing a continuously up-to-date picture of MVD epidemiological knowledge. We intend to further expand the database to other pathogens in the near future.

## Declarations

- **Funding** We acknowledge funding from the MRC Centre for Global Infectious Disease Analysis (reference MR/R015600/1) jointly funded by the UK Medical Research Council (MRC) and the UK Foreign, Commonwealth & Development Office (FCDO), under the MRC/FCDO Concordat agreement and is also part of the EDCTP2 programme supported by the European Union; the National Institute for Health Research (NIHR) for support for the Health Research Protection Unit for Modelling and Health Economics, a partnership between UK Health Security Agency, Imperial College London and London School of Hygiene & Tropical Medicine (LSHTM) (grant code NIHR200908). Additional individual funding sources: CM acknowledges the Schmidt Foundation for research funding (grant code 6-22-63345); PD acknowledges funding by Community Jameel; GCD acknowledges funding from the Royal Society; RM acknowledges the NIHR Health Protection Research Unit (HPRU) in Emerging and Zoonotic Infections, a partnership between PHE, University of Oxford, University of Liverpool and Liverpool School of Tropical Medicine (grant code NIHR200907); JW acknowledges research funding from the Wellcome Trust (grant 102169/Z/13/Z); RKN acknowledges research funding from the Medical Research Council (MRC) Doctoral Training Partnership (grant MR/N014103/1); JS acknowledges research funding from the Wellcome Trust (grant 215163/Z/18/Z); KM acknowledges research funding from the Imperial College President’s PhD Scholarship. For the purpose of open access, the author has applied a ‘Creative Commons Attribution’ (CC BY) licence to any Author Accepted Manuscript version arising from this submission.
- **Availability of data and materials** https://github.com/mrc-ide/epireview/tree/main/data
- **Code availability** https://github.com/mrc-ide/epireview
- **PROSPERO** CRD42023393345 (https://www.crd.york.ac.uk/prospero/display_record.php?RecordID=393345)
- **Competing interests** The views expressed are those of the author(s) and not necessarily those of the NIHR, UK Health Security Agency or the Department of Health and Social Care. NI is currently employed by Wellcome. However, Wellcome had no role in the the design and conduct of the study; collection, management, analysis, and interpretation of the data; preparation, review, or approval of the manuscript; and decision to submit the manuscript for publication.
- **Authors’ contributions** AC, SvE, SB, NI were responsible for conceptualisation of the study. GCD, KM, RM, HJTU, PD, RKN, JTH, KC, CG, BL, DN, JS, JW, SvE, AC, CM were responsible for data curation. GCD, KM, RM, CM were responsible for formal analysis. AC was responsible for acquisition of funding. All authors were responsible for the methodology. GCD, MK, SB, NI, SvE, AC, CM were responsible for project administration. PD, DN, MK, SvE were responsible for resources. AC, CM were responsible for supervision of the study. GCD, KM, RM, CM were responsible for validation of the data. GCD, KM, RM, CM were responsible for visualisation of the data. GCD, KM, RM, HJTU, KC, BL, AC, CM were responsible for writing the original draft of the manuscript. All authors were responsible for review and editing of the manuscript. All authors debated, discussed, edited, and approved the final version of the manuscript. GCD, KM, RM, PD, RKN, CM had full access to the full data in the study and verified all the data. All authors had final responsibility for the decision to submit the manuscript for publication. The authors alone are responsible for the views expressed in this publication and they do not necessarily represent the decisions, policy, or views of any funding bodies.

## Data Availability

All data produced are available online at https://github.com/mrc-ide/epireview

## Acknowledgements

Membership of the **Pathogen Epidemiology Review Group** (in alphabetical order by surname): Marc Baguelin (Imperial College London), Sangeeta Bhatia (Imperial College London), Lorenzo Cattarino (UKHSA), Kelly Charniga (Imperial College London), Anne Cori (Imperial College London), Zulma Cucunuba Perez (Pontificia Universidad Javeriana), Gina Cuomo-Dannenburg (Imperial College London), Amy Dighe (Johns Hopkins), Patrick Doohan (Imperial College London), Alpha Forna (University of Georgia), Keith Fraser (Imperial College London), Lily Geidelberg (), Cyril Geismar (Imperial College London), Arran Hamlet (Imperial College London), Joseph T. Hicks (Imperial College London), Natsuko Imai (Imperial College London), David Jorgensen (Imperial College London), Ed Knock (Imperial College London), Mara Kont (Imperial College London), Ben Lambert (University of Exeter), Ruth McCabe (University of Oxford), Kelly McCain (Imperial College London), Christian Morgenstern (Imperial College London), Aaron Morris (University of Oxford), Rebecca K. Nash (Imperial College London), Dariya Nikitin (Imperial College London), Sreejith Radhakrishnan (University of Glasgow), Isobel Routledge (UCSF), Janetta Skarp (Imperial College London), Hayley Thompson (PATH), H. Juliette T. Unwin (Imperial College London), Sabine van Elsland (Imperial College London), Jack Wardle (Imperial College London), Charlie Whittaker (Imperial College London)

