## Supplemental Information for "Marburg Virus Disease outbreaks, mathematical models, and disease parameters: a Systematic Review"

**B Additional Figures & Tables** **2**

**C epireview** **8**

**D PRISMA 2020 Checklists** **8**

**E Systematic Reviews** **11**

**F Excluded Studies** **11**

### A Additional Information on Methods

#### A.1 Study selection

Original research papers in English were included if reporting on MVD transmission, evolution, natural history, severity, seroprevalence, size of previous outbreaks or published mathematical transmission models. Non-peer reviewed literature was excluded. Papers identified in the search were imported into *Covidence*, a software program used to manage systematic reviews. From a team of seven reviewers, two independent reviewers first screened titles and abstracts then full texts to assess eligibility for data extraction. Disagreements in eligibility determination were resolved by consensus between the two independent reviewers.

**Search terms:** `Marburg virus AND ((transmission OR epidemiology) OR (model* NOT imag*) OR (\burden" OR \severity" OR \case fatality ratio" OR \CFR") OR (\serial interval" OR \incubation period" OR \generation time") OR (\heterogeneity" OR \superspread*") OR (\reproduction number" OR "reproductive number" OR "R0") OR (\pre-existing immunity" OR \serological" OR \serology" OR \serosurveys") OR (diagnostic OR diagnosis OR test*) OR ("evolutionary rate" OR "genetic mutation" OR evolution) OR (outbreak OR cluster OR epidemic) OR (\risk factor*") OR (\case definition"))`

#### A.2 Data Extraction

##### Outbreaks

Possible methods of case confirmation included rapid diagnostic tests (RDTs) or polymerase chain reaction (PCR) tests.

##### Models

Details of models of disease transmission were extracted, including model type, whether it modelled deterministic or stochastic processes, whether the model was theoretical or fitted to data, availability of model code, and in the case of a compartmental model, model subclassification (e.g., SIR, SEIR). Finally, we extracted information on transmission routes modelled, underlying model assumptions, and interventions included within the model.

**Parameters**

Survey context of parameter values included survey location and dates, sample size, basic demographic information, and timing of the survey in relation to reported outbreaks. For each parameter, we extracted all available information, including types of values (e.g., mean, standard deviation, median), uncertainty intervals (capturing the precision of estimates), and ranges (if when multiple estimates were obtained from different populations or using different methods).

We recorded the methods used for reproduction number estimation (e.g. renewal equations, empirical methods, compartmental models). For case fatality ratios, we extracted whether the estimation approach accounted for cases with unknown final status or not. For case fatality ratios and seroprevalence, we additionally recorded numerators and denominators where available. For genomic data, we noted the gene studied if specified and if the sequence data were available.

Outcome (e.g. infection or death), the risk factor for that outcome (e.g. age, sex or occupation), the type of occupation if specified, and whether the risk factor(s) estimates were statistically significant and/or adjusted. We chose not to extract odds ratio estimates because studies may have used different stratifications or reference groups, making it challenging to compare values across studies. The information we extract offers an overview of risk factors explored across studies that may affect the risk of infection and death, which may be useful to consider when designing MVD transmission models.

#### A.3 Analysis

We presented the de-duplicated start and end dates, deaths, confirmed, suspected, asymptomatic, and severe/hospitalised cases, and method of case confirmation in Table S2.

Confidence intervals for the unadjusted CFR estimates, as shown in Figure S2, were computed as $(\hat{CFR} \pm 1.96 \times \sqrt{\frac{\hat{CFR}\times(1-\hat{CFR})}{n}}$, where $n$ denotes the number of cases (e.g. the denominator used to derive the CFR estimate).

#### A.4 Meta-analysis methods

We provide a brief overview of the methodology used in the meta-analysis of CFRs in Figure 2. We followed a standard methodology for systematic reviews, and all analyses were performed using the `meta` R package [19]. A mixed effects model is a linear model such that $y_i = \beta_0 + \sum_j \beta_j x_i + u_i + \epsilon_i$, where $\beta = (\beta_0, ..., \beta_j)^\top$ represent the fixed effects, $y_i$ the observed data, $x_i$ any explanatory variable and $u_i$ are the random effect terms, centred around zero and independent across $i$, and $\epsilon_i$ are error terms. Meta-analysis is a special case of the above mixed-effects model with only an intercept term ($\beta_0$) and a random-effects term $u_i$ associated with that intercept.

We derived a CFR for each study, either using the CFR directly as reported in the paper or by calculating the unadjusted CFR from outbreak data (deaths/cases). We then transformed the CFRs using logit-transformation $y_i = \log\left(\frac{CFR_i}{1-CFR_i}\right)$ to ensure that the distribution was approximately normal. Finally, we used a generalised logistic mixed-effects model with the transformed CFRs as the outcome to estimate the pooled effect.

A comprehensive overview of the methodology is provided by [59].

### B Additional Figures & Tables

| Inclusion | Exclusion |
|---|---|
| Measures/estimates of human: Reproduction numbers (R, $R_0$, $R_t$, r, $R_e$), growth rate (r), doubling times, generation time, serial interval, incubation/latent period, case fatality ratio (CFR), attack rate, mutation rate (e.g. from phylogenetic study), overdispersion, risk factors (risk and the measure). | Non-English language publication |
| Mention of historical or any outbreak in humans: size, year, location, duration, spatial scale | Studies of co-infections. (local, regional, national, international). |

Table S1: Inclusion and exclusion criteria.

| **Inclusion** | **Exclusion** |
|---|---|
| Measures/estimates of animal: R, $R_0$, $R_t$, r, $R_e$, growth rate, mutation rate. | Animal studies that do not report R, $R_t$ etc. |
| Mathematical or statistical model of transmission. | Qualitative studies, e.g., KAP studies. |
| Measures of seroprevalence and negative seroprevalence in humans. | Pathogen not the primary focus of study. |
| Relative ratio of human-human vs animal introductions. | Duplicates. |
| Reviews that report inclusion criteria for reference checking. | Does not match any of the inclusion criteria. |
| For "small" pathogens, include case reports to potentially reconstruct serial interval distribution etc. | In-vitro studies. |
| | Non-peer reviewed publications (conference proceedings, abstracts, posters, letters to the editor) |
| | Papers that reference the city of Marburg in Germany instead of MVD |

Table S1: Inclusion and exclusion criteria.

| Data field | Expected data type | Variable name | Notes |
|---|---|---|---|
| Outbreak ID | integer | outbreak_id | ID assigned by database |
| Article ID | integer | article_id | ID to connect to article form |
| Outbreak start day | integer | outbreak_start_day | Day of outbreak start if reported |
| Outbreak start month | character | outbreak_start_month | Month of outbreak start if reported |
| Outbreak start year | integer | outbreak_start_year | Year of outbreak start if reported |
| Outbreak end day | integer | outbreak_end_day | Day of outbreak end if reported |
| Outbreak end month | character | outbreak_end_month | Month of outbreak end if reported |
| Outbreak end year | integer | outbreak_date_year | Year of outbreak end if reported |
| Duration (months) | integer | outbreak_duration_months | Duration of outbreak in months, if reported. No calculation of duration is done. |
| Asymptomatic transmission described | logical | asymptomatic_transmission | Tick box whether asymptomatic transmission is described in the paper or not. |
| Outbreak country | character | outbreak_country | Country or countries where the outbreak took place - from dropdown list |
| Outbreak location | character | outbreak_location | Region/district/province/city where the outbreak took place |
| Cases confirmed | integer | cases_confirmed | Number of confirmed cases as reported |
| Mode of detection of cases | character | cases_mode_detection | Method for case detection - from dropdown list |
| Cases suspected | integer | cases_suspected | Number of suspected cases as reported |
| Asymptomatic cases | integer | cases_asymptomatic | Number of asymptomatic cases as reported |
| Deaths | integer | deaths | Number of deaths as reported |

Table S2: Outbreak form fields. Refer to epireview in Supplement C for dropdown options.

| Data field | Expected data type | Variable name | Notes |
|---|---|---|---|
| Model data ID | integer | model_data_id | ID assigned by database |
| Article ID | integer | article_id | ID to connect to article form |
| Model type | character | model_type | General type of model - from dropdown list |
| Compartmental type | character | compartmental_type | Specific type of compartmental model - from dropdown list |
| Stochastic or deterministic | character | stoch_deter | Stochastic or deterministic model as reported |
| Theoretical model | logical | theoretical_model | Tick box whether the model was fitted to data (NA) or just theoretical (TRUE) |
| Intervention type | character | interventions_type | Type of intervention(s) modelled - from dropdown list |
| Code available | logical | code_available | Tick box whether code for model was publicly available and reported in the paper |
| Transmission route | character | transmission_route | Transmission route(s) modelled - from dropdown list |
| Assumptions | character | assumptions | General assumptions for the model - from dropdown list |

Table S3: Model form fields. Refer to epireview in Supplement C for dropdown options.

| Data field | Expected data type | Variable name | Notes |
|---|---|---|---|
| Parameter data ID | integer | parameter_data_id | ID assigned by database |
| Article ID | integer | article_id | ID to connect to article form |
| Parameter type | character | parameter_type | Category of parameter - see dropdown list |
| Parameter value | numeric | parameter_value | Central parameter value |
| Parameter unit | character | parameter_unit | Units for parameter value, applies to central estimate and ranges/uncertainty intervals - see dropdown list |
| Parameter lower bound | numeric | parameter_lower_bound | Lower bound of the parameter range if a range was reported or if data are disaggregated |
| Parameter upper bound | numeric | parameter_upper_bound | Upper bound of the parameter range if a range was reported or if data are disaggregated |
| Parameter value type | character | parameter_value_type | Type of central parameter value - see dropdown list |
| Parameter uncertainty - single value | numeric | parameter_uncertainty_single_value | Value for uncertainty for central parameter value if a single value was reported (e.g. value of std. dev.) |
| Parameter uncertainty - single type | character | parameter_uncertainty_singe_type | Type of uncertainty fpr central parameter value if single value was reported - see dropdown list |

| | | | |
|---|---|---|---|
| Parameter uncertainty - lower value | numeric | parameter_uncertainty_lower_value | Lower bound for uncertainty for central parameter value if paired values were reported |
| Parameter uncertainty - upper value | numeric | parameter_uncertainty_upper_value | Upper bound for uncertainty for central parameter value if paired values were reported |
| Parameter uncertainty paired type | character | parameter_uncertainty_type | Type of uncertainty for central parameter value if paired values were reported - see dropdown list |
| Numerator | integer | cfr_ifr_numerator | Numerator of either CFR/IFR (deaths) or seroprevalence (number seropositive) estimates |
| Denominator | integer | cfr_ifr_denominator | Denominator of either CFR/IFR (cases) or seroprevalence (number tested) estimates |
| Distribution type | logical | distribution_type | Type of distribution for estimated parameter - see dropdown list |
| First distribution parameter value | logical | distribution_par1_value | Value for first distribution parameter (e.g. shape or scale parameter for a gamma distribution) |
| First distribution parameter type | logical | distribution_par1_type | Type of value for first distribution parameter - see dropdown list |
| First distribution parameter uncertainty | logical | distribution_par1_uncertainty | Tick box for whether uncertainty is estimated for the first distribution parameter (TRUE) or not (FALSE) |
| Second distribution parameter value | logical | distribution_par2_value | Value for second distribution parameter (e.g. shape or scale parameter for a gamma distribution) |
| Second distribution parameter type | logical | distribution_par2_type | Type of value for second distribution parameter - see dropdown list |
| Second distribution parameter uncertainty | logical | distribution_par2_uncertainty | Tick box for whether uncertainty is estimated for the second distribution parameter (TRUE) or not (FALSE) |
| Is parameter from supplement? | logical | method_from_supplement | Tick box for whether parameter was extracted from supplement (TRUE) or not (FALSE) |
| Survey timing related to outbreak | character | method_moment_value | Timing of the survey in relation to the outbreak, if specified in paper - see dropdown list |
| Is the CFR/IFR estimate adjusted? | character | cfr_ifr_method | Is the CFR/IFR estimate adjusted, unadjusted, or unspecified - see dropdown list |
| Method to estimate R | character | method_r | Method used for estimation of the reproduction number - see dropdown list |
| Parameter estimates disaggregated by | character | method_disaggregated_by | Categories for disaggregation of parameter estimates |
| Disaggregated data available | logical | method_disaggregated | Tick box if disaggregated estimates are available (TRUE) or not (FALSE) |
| Only disaggregated data available | logical | method_disaggregated_only | Tick box if ONLY disaggregated estimates are available (TRUE) or if a central estimate is also available (FALSE) |
| Outcome for risk factor(s) | character | riskfactor_outcome | Outcome for risk factor(s) - see dropdown list |
| Risk factor name | character | riskfactor_name | Risk factor name - see dropdown list |

| | | | |
|---|---|---|---|
| Risk factor occupation | character | riskfactor_occupation | If risk factor is an occupation, then specified occupation as risk factor - see dropdown list |
| Risk factor significant | character | riskfactor_significant | Statistical significance of risk factor(s) - see dropdown list |
| Risk factor adjusted | character | riskfactor_adjusted | Adjustment status of risk factor(s)- see dropdown list |
| Sex of study population | character | population_sex | Sex of survey population - see dropdown list |
| Population sample setting | character | population_sample_type | General setting of the survey - see dropdown list |
| Population group | character | population_group | Specific group of the survey population - see dropdown list |
| Study population minimum age (years) | numeric | population_age_min | Minimum age of the survey population in years |
| Study population maximum age (years) | numeric | population_age_max | Maximum age of the survey population in years |
| Study population sample size | integer | population_sample_size | Sample size of the population used for parameter estimation |
| Study population country | character | population_country | Country of the survey population - see dropdown list |
| Study population location | character | population_location | Region/district/province/city of the survey population - see dropdown list |
| Start day of study | integer | population_study_start_day | Study start day |
| Start month of study | character | population_study_start_month | Study start month - see dropdown list |
| Start year of study | integer | population_study_start_year | Study start year - see dropdown list |
| End day of study | integer | population_study_end_day | Study end day |
| End month of study | character | population_study_end_month | Study end month - see dropdown list |
| End year of study | integer | population_study_end_year | Study end year - see dropdown list |
| Genome site | character | genome_site | Site of genome or gene studied |
| Genomic sequence available? | logical | genomic_sequence_available | Tick box whether genomic sequence data are available (TRUE) or not (FALSE) |
| Parameter class | character | parameter_class | General parameter class (delays, seroprevalence, reproduction numbers, mutations, severity, risk factors, relative contribution) |
| Uncertainty | character | Uncertainty | Formatted uncertainty range from ‘parameter_uncertainty_lower_value‘ and ‘parameter_uncertainty_lower_value‘ in format x - x for ranges and x, x for confidence or credible intervals |
| Survey year | character | Survey year | Dates of survey in format YYYY, YYYY-YYYY, MMM YYYY, or MMM-MMM YYYY from survey start and end variables |

Table S4: Parameter form fields. Refer to epireview in Supplement C for dropdown options.

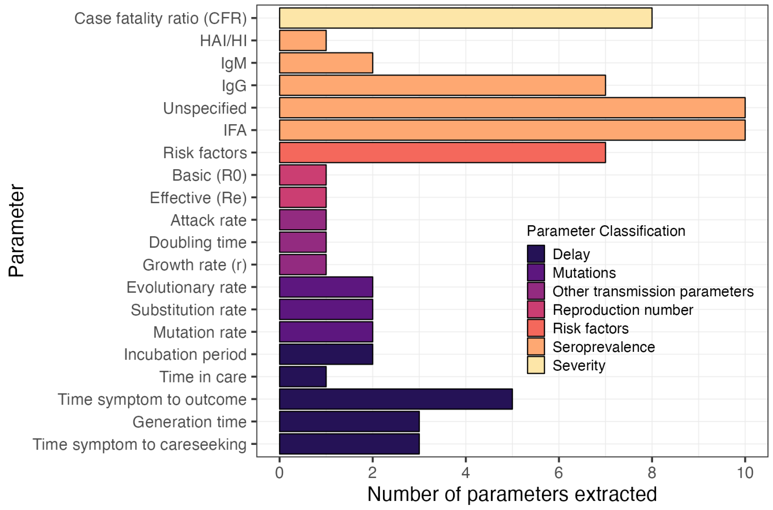

Figure S1: The number of each type of parameter extracted from studies included in the review.

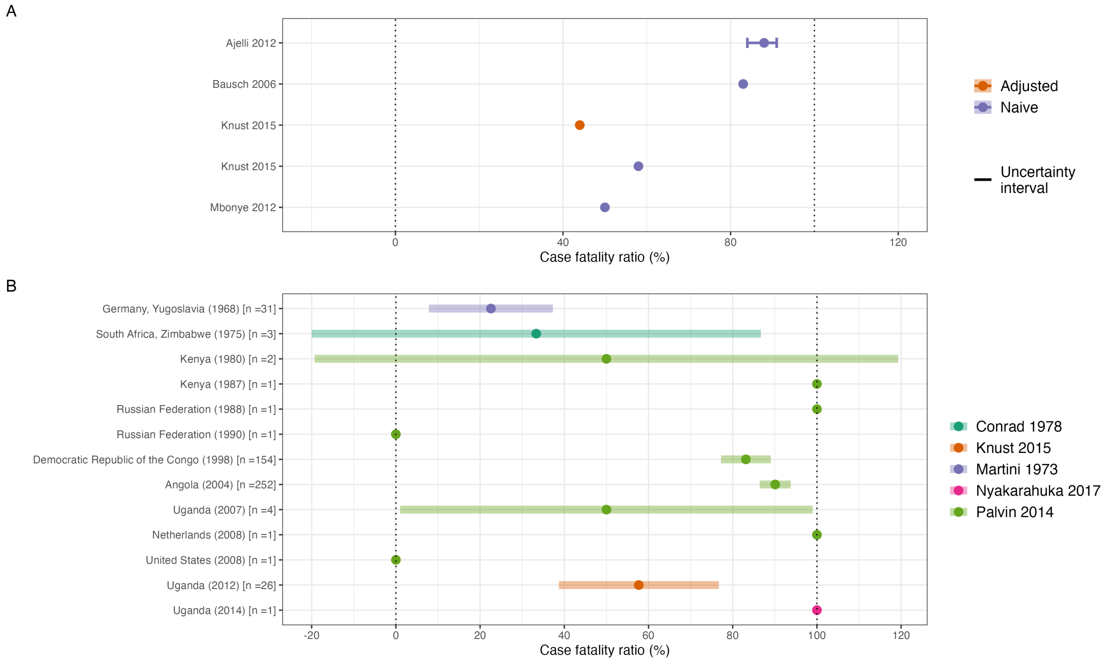

Figure S2: Overview of the estimates of the case fatality ratio (CFR) obtained from the included studies. (A) CFR estimates reported in the included studies, stratified according to estimation method. Points represent central estimates. Error bars represent an uncertainty interval associated with the point estimate, as reported in the original study. (B) CFR estimated from extracted outbreak data, including only one observation per outbreak using the study with the longest duration of the outbreak reported ensuring each case is not double counted. Shaded bars represents the imputed binomial confidence interval for studies with a sample size, $n > 1$. Vertical dotted lines represent 0% and 100% CFR.

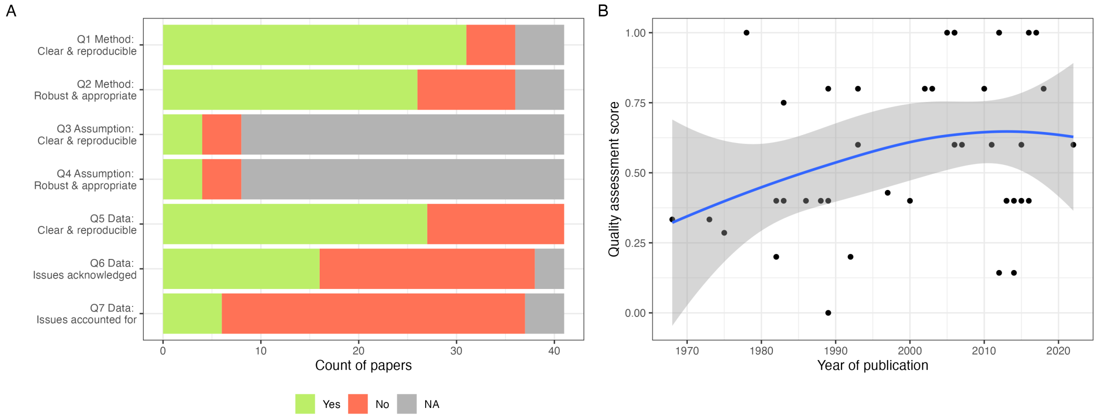

Figure S3: (A) Count of papers for each quality assessment question scoring Yes, No or not applicable. (B) Quality Assessment Score defined as proportion of Yes votes for each paper relative to sum of Yes and No answers, removing NAs. The time trend is fitted using Local Polynomial Regression Fitting.

| Article | Country | Survey year | Outcome | Risk factor | Significant | Adjusted | Sample size | Population sample type | Population group | Timing of survey |
|---|---|---|---|---|---|---|---|---|---|---|
| Borchert 2005 | Angola | Mar-Jul 2005 | Infection | Gathering | Significant | Unknown | 102 | Hospital based | Persons under investigation | Mid outbreak |
| Borchert 2005 | Angola | Mar-Jul 2005 | Infection | Occupation - Funeral and burial services | Significant | Unknown | 102 | Hospital based | Persons under investigation | Mid outbreak |
| Borchert 2005 | Angola | Mar-Jul 2005 | Infection | Household contact | Significant | Unknown | 102 | Hospital based | Persons under investigation | Mid outbreak |
| Borchert 2005 | Angola | Mar-Jul 2005 | Infection | Other | Significant | Unknown | 102 | Hospital based | Persons under investigation | Mid outbreak |
| Knust 2015 | Uganda | 2012 | Infection | Gathering | Significant | Unknown | 26 | Community based | General population | Post outbreak |
| Knust 2015 | Uganda | 2012 | Infection | Other | Not significant | Unknown | 26 | Community based | General population | Post outbreak |
| Knust 2015 | Uganda | 2012 | Infection | Sex | Not significant | Unknown | 26 | Community based | General population | Post outbreak |
| Amman 2012 | Uganda | | Infection | Contact with animal | Significant | Unknown | | | | |
| Amman 2012 | Uganda | | Infection | Other | Significant | Unknown | | | | |
| Bausch 2003 | Democratic Republic of the Congo | May 1999 | Serology | Hospitalisation | Significant | Adjusted | 915 | Population based | | |
| Bausch 2003 | Democratic Republic of the Congo | May 1999 | Serology | Contact with animal | Not significant | Adjusted | 912 | Population based | | |
| Bausch 2003 | Democratic Republic of the Congo | May 1999 | Serology | Household contact | Not significant | Adjusted | 912 | Population based | | |
| Bausch 2003 | Democratic Republic of the Congo | May 1999 | Serology | Occupation - Funeral and burial services | Not significant | Adjusted | 912 | Population based | | |
| Borchert 2005 | Democratic Republic of the Congo | | Serology | Contact with animal | Not significant | Unknown | 300 | Community based | | Post outbreak |
| Borchert 2005 | Democratic Republic of the Congo | | Serology | Gathering | Not significant | Unknown | 300 | Community based | | Post outbreak |

Figure S4: More detailed table of risk factor data from the extracted studies, giving countries, times and contexts of surveys and non-aggregated information on each risk factor assessed in the four relevant studies.

### C epireview

We developed an R package called *epireview* that provides a central location to host and access the extracted data for the nine priority pathogens, allows for submissions of outbreak, model, and parameter data from new peer-reviewed papers via pull requests, and includes functions to produce the figures and tables included in this paper and update them with any additional data. This package will be updated as the overall project by the Pathogen Epidemiology Review Group (PERG) continues to extract modelling parameters for the rest of the nine priority pathogens as defined by WHO.

There are several vignettes available:

- A vignette for MVD with tables and figures from this paper that will be updated as data are added to the database.
- A vignette that lists the options for each model, outbreak, or parameter field and describes how to access them using a function in the package.
- A vignette to explain the process of updating the database with new article, model, outbreak, or pathogen data.

### D PRISMA 2020 Checklists

| Section & Topic | Item # | Checklist item | Reported (Yes/No) |
|---|---|---|---|
| **Title** | | | |
| Title | 1 | Identify the report as a systematic review. | Yes |
| **Background** | | | |
| Objectives | 2 | Provide an explicit statement of the main objective(s) or question(s) the review addresses. | Yes |
| **Methods** | | | |
| Eligibility criteria | 3 | Specify the inclusion and exclusion criteria for the review. | Yes |
| Information sources | 4 | Specify the information sources (e.g. databases, registers) used to identify studies and the date when each was last searched. | Yes |
| Risk of bias | 5 | Specify the methods used to assess risk of bias in the included studies. | Yes |
| Synthesis of results | 6 | Specify the methods used to present and synthesise results. | Yes |
| **Results** | | | |
| Included studies | 7 | Give the total number of included studies and participants and summarise relevant characteristics of studies. | Yes |
| Synthesis of results | 8 | Present results for main outcomes, preferably indicating the number of included studies and participants for each. If meta-analysis was done, report the summary estimate and confidence/credible interval. If comparing groups, indicate the direction of the effect (i.e. which group is favoured). | Yes |
| **Discussion** | | | |
| Limitations of evidence | 9 | Provide a brief summary of the limitations of the evidence included in the review (e.g. study risk of bias, inconsistency and imprecision). | Yes |
| Interpretation | 10 | Provide a general interpretation of the results and important implications. | Yes |
| **Other** | | | |
| Funding | 11 | Specify the primary source of funding for the review. | Yes |
| Registration | 12 | Provide the register name and registration number. | Yes |

Table S5: PRISMA 2020 Abstracts Checklist. ([60])

| Section & Topic | Item # | Checklist item | Location where item is reported |
|---|---|---|---|
| **Title** | | | |
| Title | 1 | Identify the report as a systematic review. | page 1 |
| **Abstract** | | | |
| Abstract | 2 | See the PRISMA 2020 for Abstracts checklist. | Table S5 |
| **Introduction** | | | |
| Rationale | 3 | Describe the rationale for the review in the context of existing knowledge. | page 2 |
| Objectives | 4 | Provide an explicit statement of the objective(s) or question(s) the review addresses. | page 2 |
| **Methods** | | | |
| Eligibility criteria | 5 | Specify the inclusion and exclusion criteria for the review and how studies were grouped for the syntheses. | page 3 |
| Information sources | 6 | Specify all databases, registers, websites, organisations, reference lists and other sources searched or consulted to identify studies. Specify the date when each source was last searched or consulted. | page 3 |
| Search strategy | 7 | Present the full search strategies for all databases, registers and websites, including any filters and limits used. | page 3 + Figure 1 |
| Selection process | 8 | Specify the methods used to decide whether a study met the inclusion criteria of the review, including how many reviewers screened each record and each report retrieved, whether they worked independently, and if applicable, details of automation tools used in the process. | page 3 |

Table S6: PRISMA 2020 Checklist. ([60])

| Section & Topic | Item # | Checklist item | Location where item is reported |
|---|---|---|---|
| Data collection process | 9 | Specify the methods used to collect data from reports, including how many reviewers collected data from each report, whether they worked independently, any processes for obtaining or confirming data from study investigators, and if applicable, details of automation tools used in the process. | page 3 |
| Data items | 10a | List and define all outcomes for which data were sought. Specify whether all results that were compatible with each outcome domain in each study were sought (e.g. for all measures, time points, analyses), and if not, the methods used to decide which results to collect. | page 3 |
| | 10b | List and define all other variables for which data were sought (e.g. participant and intervention characteristics, funding sources). Describe any assumptions made about any missing or unclear information. | page 3 |
| Study risk of bias assessment | 11 | Specify the methods used to assess risk of bias in the included studies, including details of the tool(s) used, how many reviewers assessed each study and whether they worked independently, and if applicable, details of automation tools used in the process. | page 3/4 |
| Effect measures | 12 | Specify for each outcome the effect measure(s) (e.g. risk ratio, mean difference) used in the synthesis or presentation of results. | page 3/4 |
| Synthesis methods | 13a | Describe the processes used to decide which studies were eligible for each synthesis (e.g. tabulating the study intervention characteristics and comparing against the planned groups for each synthesis (item 5)). | page 3/4 |
| | 13b | Describe any methods required to prepare the data for presentation or synthesis, such as handling of missing summary statistics, or data conversions. | page 3/4 |
| | 13c | Describe any methods used to tabulate or visually display results of individual studies and syntheses. | page 3/4 |
| | 13d | Describe any methods used to synthesize results and provide a rationale for the choice(s). If meta-analysis was performed, describe the model(s), method(s) to identify the presence and extent of statistical heterogeneity, and software package(s) used. | page 3/4 |
| | 13e | Describe any methods used to explore possible causes of heterogeneity among study results (e.g. subgroup analysis, meta-regression). | - |
| | 13f | Describe any sensitivity analyses conducted to assess robustness of the synthesized results. | - |
| Reporting bias assessment | 14 | Describe any methods used to assess risk of bias due to missing results in a synthesis (arising from reporting biases). | page 4 |
| Certainty assessment | 15 | Describe any methods used to assess certainty (or confidence) in the body of evidence for an outcome. | page 4 |
| **Results** | | | |
| Study selection | 16a | Describe the results of the search and selection process, from the number of records identified in the search to the number of studies included in the review, ideally using a flow diagram. | page 4 |
| | 16b | Cite studies that might appear to meet the inclusion criteria, but which were excluded, and explain why they were excluded. | Table S8 |
| Study characteristics | 17 | Cite each included study and present its characteristics. | pages 5-13 |
| Risk of bias in studies | 18 | Present assessments of risk of bias for each included study. | page 13 |
| Results of individual studies | 19 | For all outcomes, present, for each study: (a) summary statistics for each group (where appropriate) and (b) an effect estimate and its precision (e.g. confidence/credible interval), ideally using structured tables or plots. | pages 5-13 |
| Results of syntheses | 20a | For each synthesis, briefly summarise the characteristics and risk of bias among contributing studies. | page 7/8 |
| | 20b | Present results of all statistical syntheses conducted. If meta-analysis was done, present for each the summary estimate and its precision (e.g. confidence/credible interval) and measures of statistical heterogeneity. If comparing groups, describe the direction of the effect. | page 7/8 |
| | 20c | Present results of all investigations of possible causes of heterogeneity among study results. | page 7/8 |
| | 20d | Present results of all sensitivity analyses conducted to assess the robustness of the synthesized results. | page 7/8 |
| Reporting biases | 21 | Present assessments of risk of bias due to missing results (arising from reporting biases) for each synthesis assessed. | page 7/8 |
| Certainty of evidence | 22 | Present assessments of certainty (or confidence) in the body of evidence for each outcome assessed. | pages 5-13 |

Table S6: PRISMA 2020 Checklist. ([60])

| Section & Topic | Item # | Checklist item | Location where item is reported |
|---|---|---|---|
| **Discussion** | | | |
| Discussion | 23a | Provide a general interpretation of the results in the context of other evidence. | page 13 |
| | 23b | Discuss any limitations of the evidence included in the review. | page 13-14 |
| | 23c | Discuss any limitations of the review processes used. | page 13-14 |
| | 23d | Discuss implications of the results for practice, policy, and future research. | page 13-14 |
| **Other Information** | | | |
| Registration and protocol | 24a | Provide registration information for the review, including register name and registration number, or state that the review was not registered. | page 14 |
| | 24b | Indicate where the review protocol can be accessed, or state that a protocol was not prepared. | page 14 |
| | 24c | Describe and explain any amendments to information provided at registration or in the protocol. | n/a |
| Support | 25 | Describe sources of financial or non-financial support for the review, and the role of the funders or sponsors in the review. | page 14 |
| Competing interests | 26 | Declare any competing interests of review authors. | page 14 |
| Availability of data, code and other materials | 27 | Report which of the following are publicly available and where they can be found: template data collection forms; data extracted from included studies; data used for all analyses; analytic code; any other materials used in the review. | page 14 |

Table S6: PRISMA 2020 Checklist. ([60])

### E Systematic Reviews

Systematic reviews were excluded from our analysis, but Table S7 presents the systematic reviews which we have found and used for validation purposes.

| Study | Title | Journal | DOI |
|---|---|---|---|
| Selvaraj 2018 | Infection Rates and Risk Factors for Infection Among Health Workers During Ebola and Marburg Virus Outbreaks: A Systematic Review | The Journal of infectious diseases | 10.1093/infdis/jiy435 |
| Brainard 2016 | Presence and Persistence of Ebola or Marburg Virus in Patients and Survivors: A Rapid Systematic Review | PLoS neglected tropical diseases | 10.1371/journal.pntd.0004475 |
| Brainard 2016 | Risk factors for transmission of Ebola or Marburg virus disease: a systematic review and meta-analysis | International journal of epidemiology | 10.1093/ije/dyv307 |
| Tahmo 2023 | An epidemiological synthesis of emerging and re-emerging zoonotic disease threats in Cameroon, 2000-2022: a systematic review | IJID Reg | 10.1016/j.ijregi.2022.12.001 |
| Nyakarahuka 2016 | How severe and prevalent are Ebola and Marburg viruses? A systematic review and meta-analysis of the case fatality rates and seroprevalence | BMC infectious diseases | 10.1186/s12879-016-2045-6 |

Table S7: Excluded systematic reviews

### F Excluded Studies

We list all excluded studies with reasons for exclusion in Table S8

| Study | Title | Journal | Notes |
|---|---|---|---|
| Zeller 2000 | Infections by viruses of the families Bunyaviridae and Filoviridae | Revue scientifique et technique (International Office of Epizootics) | Exclusion reason: Reports metrics from other papers (not original estimates of primary data); JS (2019-03-21 21:30:42)(Screen): mention of historical outbreaks; |
| Wei 2017 | Deep-sequencing of Marburg virus genome during sequential mouse passaging and cell-culture adaptation reveals extensive changes over time | Scientific reports | Exclusion reason: No report of parameters (including seroprevalence and other measures of interest) or transmission models or historical outbreaks; JS (2019-08-08 23:45:59)(Select): Yes, maybe someone with knowledge of genomics could calculate a mutation rate out of info here..?; KC (2019-03-19 23:05:13)(Screen): mutation rate?; |
| VanKerkhove 2015 | A review of epidemiological parameters from Ebola outbreaks to inform early public health decision-making | Scientific data | Exclusion reason: Reports metrics from other papers (not original estimates of primary data); |
| Tukei 1996 | Threat of Marburg and Ebola viral haemorrhagic fevers in Africa | East African medical journal | Exclusion reason: Reports metrics from other papers (not original estimates of primary data); |
| Sweileh 2017 | Global research trends of World Health Organization's top eight emerging pathogens | Globalization and health | Exclusion reason: Wrong pathogen or pathogen epidemiology or transmission not main focus; JS (2019-08-08 19:54:12)(Select): Nothing super relevant transmission-wise though; JS (2019-08-08 19:53:58)(Select): Could check if we have similar split of disease papers as them; JS (2019-03-20 21:48:23)(Screen): massive review - good for reference checking; |
| Swanepoel 2007 | Studies of reservoir hosts for Marburg virus | Emerging infectious diseases | Exclusion reason: No report of parameters (including seroprevalence and other measures of interest) or transmission models or historical outbreaks; JS (2019-08-08 19:49:49)(Select): Can we get mutation rate from the phylogenetic tree?; JS (2019-03-20 21:46:18)(Screen): mention of outbreak in humans; |
| Strickland-Cholmley 1970 | Marburg virus | Lancet (London, England) | Exclusion reason: No report of parameters (including seroprevalence and other measures of interest) or transmission models or historical outbreaks; |
| Storm 2018 | Antibody Responses to Marburg Virus in Egyptian Rousette Bats and Their Role in Protection against Infection | Viruses | Exclusion reason: No report of parameters (including seroprevalence and other measures of interest) or transmission models or historical outbreaks; JS (2019-08-08 19:41:03)(Select): There was some idea of infectious time, but not clearly outlined. Therefore not adding in ; JS (2019-03-13 04:22:07)(Screen): May contain data that could be used to estimate length of animal infectious period; |
| Spence 1982 | Marburg virus disease–an indicator case in South Africa | South African medical journal = Suid-Afrikaanse tydskrif vir geneeskunde | Exclusion reason: No report of parameters (including seroprevalence and other measures of interest) or transmission models or historical outbreaks; JS (2019-08-09 18:39:34)(Select): correspondence, not a paper; |
| Snowden 1979 | Marburg disease: the 20th Century | Zimbabwe Rhodesia Nurse | Exclusion reason: Full text not found; |
| Slenczka 2007 | Forty years of marburg virus | The Journal of infectious diseases | Exclusion reason: Duplicate; |
| Slenczka 1999 | The Marburg virus outbreak of 1967 and subsequent episodes | Current topics in microbiology and immunology | Exclusion reason: Reports metrics from other papers (not original estimates of primary data); JS (2019-08-23 18:53:35)(Select): Disease duration data (Table 4) - not sure what paper this comes from, also a table on secondary cases (possibly for attack rate - Table 3); JS (2019-03-20 21:24:52)(Screen): References Marburg outbreaks; |

Table S8: Excluded studies at full text review with exclusion reason

| Study | Title | Journal | Notes |
|---|---|---|---|
| Slenczka 2017 | Filovirus Research: How it Began | Current topics in microbiology and immunology | Exclusion reason: Not peer-reviewed paper; JS (2019-07-09 21:22:21)(Select): Book chapter; JS (2019-03-20 21:24:30)(Screen): mention of outbreak; |
| Siya 2019 | Lowland grazing and Marburg virus disease (MVD) outbreak in Kween district, Eastern Uganda | BMC public health | Exclusion reason: No report of parameters (including seroprevalence and other measures of interest) or transmission models or historical outbreaks; JS (2019-08-07 23:38:17)(Select): collected opinions from people, no numbers; |
| Selvaraj 2018 | Infection Rates and Risk Factors for Infection Among Health Workers During Ebola and Marburg Virus Outbreaks: A Systematic Review | The Journal of infectious diseases | Exclusion reason: Reports metrics from other papers (not original estimates of primary data); JS (2019-08-07 23:18:44)(Select): Has papers on risk factors; |
| Saluzzo 1981 | [Antibodies against the Marburg virus among human populations in the southeastern Central African Republic] | Comptes rendus des seances de l'Academie des sciences. Serie III, Sciences de la vie | Exclusion reason: Not in English; |
| Rougeron 2015 | Ebola and Marburg haemorrhagic fever | Journal of clinical virology : the official publication of the Pan American Society for Clinical Virology | Exclusion reason: Reports metrics from other papers (not original estimates of primary data); JS (2019-08-07 23:04:17)(Select): Has a list of outbreaks, but not much else; JS (2019-03-20 20:50:36)(Screen): Review - may have useful references; |
| Raabea 2012 | Infection control during filoviral hemorrhagic Fever outbreaks | Journal of global infectious diseases | Exclusion reason: No report of parameters (including seroprevalence and other measures of interest) or transmission models or historical outbreaks; |
| Polonsky 2014 | Emerging filoviral disease in Uganda: proposed explanations and research directions | The American journal of tropical medicine and hygiene | Exclusion reason: No report of parameters (including seroprevalence and other measures of interest) or transmission models or historical outbreaks; JS (2019-08-05 22:56:51)(Select): About Marburg but does not report parameters of interest; JS (2019-07-09 21:04:42)(Select): Perspectives; JS (2019-03-20 19:41:20)(Screen): mentions outbreaks, may contain useful references for the outbreaks; |
| Pittalis 2009 | Case definition for Ebola and Marburg haemorrhagic fevers: a complex challenge for epidemiologists and clinicians | The new microbiologica | Exclusion reason: No report of parameters (including seroprevalence and other measures of interest) or transmission models or historical outbreaks; JS (2019-08-05 22:38:13)(Select): The table with case definition will help fill out the summary table, they have summarised what was considered a case in many outbreaks ; JS (2019-03-20 19:31:08)(Screen): may have data on incubation periods and so on; |
| Pigott 2015 | Mapping the zoonotic niche of Marburg virus disease in Africa | Transactions of the Royal Society of Tropical Medicine and Hygiene | Exclusion reason: No report of parameters (including seroprevalence and other measures of interest) or transmission models or historical outbreaks; JS (2019-08-02 21:08:04)(Select): Not sure if this model is exactly what we want though; JS (2019-03-20 03:02:08)(Screen): modelling transmission; |
| Peterson 2016 | Geographic potential of disease caused by Ebola and Marburg viruses in Africa | Acta tropica | Exclusion reason: No report of parameters (including seroprevalence and other measures of interest) or transmission models or historical outbreaks; JS (2019-08-01 23:21:41)(Select): Nevermind, the whole paper is about reservoir distribution, not really transmission. Therefore, the pathogen epidemiology and transmission aren't really a focus here.; JS (2019-03-20 19:27:36)(Screen): model of disease transmission; |

Table S8: Excluded studies at full text review with exclusion reason

| Study | Title | Journal | Notes |
|---|---|---|---|
| Peterson 2006 | Geographic potential for outbreaks of Marburg hemorrhagic fever | The American journal of tropical medicine and hygiene | Exclusion reason: No report of parameters (including seroprevalence and other measures of interest) or transmission models or historical outbreaks; |
| Peterson 2004 | Potential mammalian filovirus reservoirs | Emerging infectious diseases | Exclusion reason: No report of parameters (including seroprevalence and other measures of interest) or transmission models or historical outbreaks; JS (2019-07-09 21:01:22)(Select): Perspective; |
| Peterson 2004 | Ecologic and geographic distribution of filovirus disease | Emerging infectious diseases | Exclusion reason: No report of parameters (including seroprevalence and other measures of interest) or transmission models or historical outbreaks; JS (2019-08-01 01:25:58)(Select): Not really transmission modelling though - more like ecologic niche modelling; JS (2019-08-01 01:18:14)(Select): Has a map of the geographic distribution of Marburg; JS (2019-03-20 19:23:45)(Screen): possibly considers transmission in the model, if not, gives an estimate of the range of occurrence of Marburg which could be interesting for considering where the outbreaks have taken place; |
| Vella 1978 | Lassa fever (LF) and Marburg disease (MVD): occurrences, origins and diagnoses | Royal Society of Health journal | Exclusion reason: No report of parameters (including seroprevalence and other measures of interest) or transmission models or historical outbreaks; JS (2019-03-21 20:19:00)(Screen): might be relevant (occurrences =outbreaks?) though paper will be hard to find; |
| Nyakarahuka 2017 | Knowledge and attitude towards Ebola and Marburg virus diseases in Uganda using quantitative and participatory epidemiology techniques | PLoS neglected tropical diseases | Exclusion reason: No report of parameters (including seroprevalence and other measures of interest) or transmission models or historical outbreaks; |
| Ndayimirije 2005 | Marburg hemorrhagic fever in Angola–fighting fear and a lethal pathogen | The New England journal of medicine | Exclusion reason: Not peer-reviewed paper; JS (2019-07-31 21:30:23)(Select): Also, no references.; JS (2019-07-09 20:47:19)(Select): Perspective piece - should be excluded?; |
| Natesan 2016 | Human Survivors of Disease Outbreaks Caused by Ebola or Marburg Virus Exhibit Cross-Reactive and Long-Lived Antibody Responses | Clinical and vaccine immunology : CVI | Exclusion reason: No report of parameters (including seroprevalence and other measures of interest) or transmission models or historical outbreaks; JS (2019-03-19 23:23:35)(Screen): might be an interesting paper regarding duration of infectiousness; |
| Miraglia 2019 | Marburgviruses: An Update | Laboratory medicine | Exclusion reason: Reports metrics from other papers (not original estimates of primary data); JS (2019-03-19 22:51:05)(Screen): Possibly useful review paper; |
| Mayega 2013 | A descriptive overview of the burden, distribution and characteristics of epidemics in Uganda | East African journal of public health | Exclusion reason: Full text not found; JS (2019-12-12 00:17:22)(Select): Application rejected by supplier; JS (2019-10-18 02:13:35)(Select): Requested from library 17/10/2019; |
| Mahanty 2004 | Pathogenesis of filoviral haemorrhagic fevers | The Lancet. Infectious diseases | Exclusion reason: Wrong pathogen or pathogen epidemiology or transmission not main focus; JS (2019-07-31 00:43:23)(Select): This paper is mostly about the cellular level response to Marburg, only thing of interest is that outbreak table but we would be getting that same table from more relevant reviews anyways; |
| MacNeil 2012 | Ebola and Marburg hemorrhagic fevers: neglected tropical diseases? | PLoS neglected tropical diseases | Exclusion reason: Reports metrics from other papers (not original estimates of primary data); |

Table S8: Excluded studies at full text review with exclusion reason

| Study | Title | Journal | Notes |
|---|---|---|---|
| Roddy 2007 | The Medecins Sans Frontieres intervention in the Marburg hemorrhagic fever epidemic, Uige, Angola, 2005. II. lessons learned in the community | The Journal of infectious diseases | Exclusion reason: Wrong pathogen or pathogen epidemiology or transmission not main focus; JS (2019-08-07 21:16:33)(Select): Has an incubation period though they do not say where they get it from; JS (2019-03-20 19:57:44)(Screen): mention of outbreak; |
| Ligon 2005 | Outbreak of Marburg hemorrhagic fever in Angola: a review of the history of the disease and its biological aspects | Seminars in pediatric infectious diseases | Exclusion reason: Reports metrics from other papers (not original estimates of primary data); |
| Leroy 2011 | Ebola and Marburg haemorrhagic fever viruses: major scientific advances, but a relatively minor public health threat for Africa | Clinical microbiology and infection : the official publication of the European Society of Clinical Microbiology and Infectious Diseases | Exclusion reason: Reports metrics from other papers (not original estimates of primary data); JS (2019-03-19 21:26:24)(Screen): references outbreaks; |
| Leffel 2004 | Marburg and Ebola viruses as aerosol threats | Biosecurity and bioterrorism : biodefense strategy, practice, and science | Exclusion reason: Reports metrics from other papers (not original estimates of primary data); |
| LeDuc 1989 | Epidemiology of hemorrhagic fever viruses | Reviews of infectious diseases | Exclusion reason: No report of parameters (including seroprevalence and other measures of interest) or transmission models or historical outbreaks; |
| Lawrence 2005 | Largest ever Marburg haemorrhagic fever outbreak, Angola | Euro surveillance : bulletin Europeen sur les maladies transmissibles = European communicable disease bulletin | Exclusion reason: Case report or case study (i.e. reports on less than 10 cases, but this threshold can be pathogen-dependent); |
| Kuzmin 2010 | Marburg virus in fruit bat, Kenya | Emerging infectious diseases | Exclusion reason: No report of parameters (including seroprevalence and other measures of interest) or transmission models or historical outbreaks; JS (2019-07-09 18:24:16)(Select): Letter to Editor; JS (2019-03-19 21:20:04)(Screen): seroprevalence in bats?; |
| Kuroda 2014 | A polymorphism of the TIM-1 IgV domain: implications for the susceptibility to filovirus infection | Biochemical and biophysical research communications | Exclusion reason: No report of parameters (including seroprevalence and other measures of interest) or transmission models or historical outbreaks; JS (2019-03-13 04:13:23)(Screen): Mentions a mortality rate so this paper may contain a reference to a paper that has CFR ; |
| Kortepeter 2011 | Basic clinical and laboratory features of filoviral hemorrhagic fever | The Journal of infectious diseases | Exclusion reason: Reports metrics from other papers (not original estimates of primary data); JS (2019-07-30 19:06:07)(Select): references for incubation period, CFR; |
| Klenk 2017 | Marburg- and Ebolaviruses: A Look Back and Lessons for the Future | Methods in molecular biology (Clifton, N.J.) | Exclusion reason: Not peer-reviewed paper; JS (2019-07-09 18:15:39)(Select): Book chapter; JS (2019-03-19 20:50:51)(Screen): has a list of Marburg outbreaks; |
| "Kalter 1969 | Antibodies in primates to the Marburg virus | Proceedings of the Society for Experimental Biology and Medicine. Society for Experimental Biology and Medicine (New York, N.Y.) | Exclusion reason: No report of parameters (including seroprevalence and other measures of interest) or transmission models or historical outbreaks; JS (2019-08-23 18:46:38)(Select): looks like negative seroprevalence in humans; JS (2019-07-09 18:10:20)(Select): Available from ICL library; JS (2019-03-13 04:10:26)(Screen): Can possibly involve humans - humans are primates ; |
| " Hayman 2015 | Biannual birth pulses allow filoviruses to persist in bat populations | Proceedings. Biological sciences | Exclusion reason: No report of parameters (including seroprevalence and other measures of interest) or transmission models or historical outbreaks; JS (2019-03-15 00:11:50)(Screen): a mathematical model of bat-human transmission?; |

Table S8: Excluded studies at full text review with exclusion reason

| Study | Title | Journal | Notes |
|---|---|---|---|
| Hartman 2010 | Ebola and marburg hemorrhagic fever | Clinics in laboratory medicine | Exclusion reason: Reports metrics from other papers (not original estimates of primary data); |
| Grolla 2011 | The use of a mobile laboratory unit in support of patient management and epidemiological surveillance during the 2005 Marburg Outbreak in Angola | PLoS neglected tropical diseases | Exclusion reason: Wrong pathogen or pathogen epidemiology or transmission not main focus; JS (2019-07-30 00:48:51)(Select): More about the performance of the laboratory than epidemiology and transmission; |
| Green 2012 | Uganda battles Marburg fever outbreak | Lancet | Exclusion reason: Case report or case study (i.e. reports on less than 10 cases, but this threshold can be pathogen-dependent); |
| Glaze 2015 | A Comparison of the Pathogenesis of Marburg Virus Disease in Humans and Nonhuman Primates and Evaluation of the Suitability of These Animal Models for Predicting Clinical Efficacy under the 'Animal Rule' | Comparative medicine | Exclusion reason: Reports metrics from other papers (not original estimates of primary data); JS (2019-07-22 23:27:49)(Select): This review has summarised Marburg outbreaks and has lots of references; JS (2019-03-19 00:55:16)(Screen): Review may have details on R0 etc.; |
| Gilsdorf 2012 | Guidance for contact tracing of cases of Lassa fever, Ebola or Marburg haemorrhagic fever on an airplane: results of a European expert consultation | BMC public health | Exclusion reason: Reports metrics from other papers (not original estimates of primary data); JS (2019-07-22 23:22:18)(Select): mentions incubation periods and historical outbreaks, and has a handy table; |
| Geisbert 2015 | Considerations in the Use of Nonhuman Primate Models of Ebola Virus and Marburg Virus Infection | The Journal of infectious diseases | Exclusion reason: Reports metrics from other papers (not original estimates of primary data); JS (2019-03-19 03:09:08)(Screen): Mentions case-fatality rates in humans - probably not the original source paper though; |
| Gear 1975 | Outbreake of Marburg virus disease in Johannesburg | British medical journal | Exclusion reason: Duplicate; |
| Gear 1989 | Clinical aspects of African viral hemorrhagic fevers | Reviews of infectious diseases | Exclusion reason: Reports metrics from other papers (not original estimates of primary data); JS (2019-07-22 21:31:03)(Select): does not report any parameters of interest of Marburg and we have the original paper on the Jburg outbreak; |
| Gear 1982 | The hemorrhagic fevers of Southern Africa with special reference to studies in the South African Institute for Medical Research | The Yale journal of biology and medicine | Exclusion reason: Reports metrics from other papers (not original estimates of primary data); JS (2019-07-22 19:45:30)(Select): Review-type article but no references and not much Marburg; |
| Galbraith 1980 | Changing patterns of communicable disease in England and Wales. Part i–Newly recognised diseases | British medical journal | Exclusion reason: Reports metrics from other papers (not original estimates of primary data); JS (2019-07-22 19:02:16)(Select): Doesn't have references for Marburg? Also, does not deal with Marburg on its own in a meaningful way; |
| Fisher-Hoch 2005 | Lessons from nosocomial viral haemorrhagic fever outbreaks | British medical bulletin | Exclusion reason: Wrong pathogen or pathogen epidemiology or transmission not main focus; JS (2019-07-19 22:54:00)(Select): Not really about Marburg on its own; |

Table S8: Excluded studies at full text review with exclusion reason

| Study | Title | Journal | Notes |
|---|---|---|---|
| Fernando 2015 | Immune Response to Marburg Virus Angola Infection in Nonhuman Primates | The Journal of infectious diseases | Exclusion reason: No report of parameters (including seroprevalence and other measures of interest) or transmission models or historical outbreaks; JS (2019-03-18 20:56:47)(Screen): Historical outbreak mentioned + CFR mentioned only in abstract/intro, not the point of the study; |
| Feldmann 1996 | Emerging and reemerging of filoviruses | Archives of virology. Supplementum | Exclusion reason: Duplicate; |
| Feldmann 1996 | Marburg and Ebola viruses | Advances in virus research | Exclusion reason: Reports metrics from other papers (not original estimates of primary data); JS (2019-08-14 22:25:32)(Select): A pretty old review, nothing here that the newer reviews would not cover. I think we can safely exclude.; |
| Feldmann 1996 | Filoviruses | Medical Microbiology | Exclusion reason: Not peer-reviewed paper; JS (2019-06-27 22:54:08)(Select): Book chapter; |
| Ewers 2016 | Natural History of Aerosol Exposure with Marburg Virus in Rhesus Macaques | Viruses | Exclusion reason: Wrong pathogen or pathogen epidemiology or transmission not main focus; PD (2023-03-14 02:23:55)(Select): Moved back to exclusion because it's an animal challenge trial; JS (2019-03-18 20:48:01)(Screen): Incubation period in animals - we care about this for animals, right? But at the same time it is an experimental study...; |
| Enserink 2005 | Infectious diseases. A puzzling outbreak of Marburg disease | Science | Exclusion reason: Not peer-reviewed paper; JS (2019-06-27 22:43:23)(Select): news - not peer-reviewed; |
| Emanuel 2018 | Filoviruses: Ecology, Molecular Biology, and Evolution | Advances in virus research | Exclusion reason: Not peer-reviewed paper; JS (2019-06-27 22:39:31)(Select): Book chapter - not a paper; |
| Dowdle 1976 | Marburg virus | Bulletin of the Pan American Health Organization | Exclusion reason: Reports metrics from other papers (not original estimates of primary data); |
| Dimitrov 2008 | Adaptive modeling of viral diseases in bats with a focus on rabies | Journal of theoretical biology | Exclusion reason: Wrong pathogen or pathogen epidemiology or transmission not main focus; JS (2019-07-18 19:05:20)(Select): The model is about bat rabies, not Marburg; JS (2019-03-15 21:56:23)(Screen): Spillover modelling; |
| Curtis 2006 | Viral haemorrhagic fevers caused by Lassa, Ebola and Marburg viruses | Advances in experimental medicine and biology | Exclusion reason: Not peer-reviewed paper; JS (2019-06-27 22:32:11)(Select): Conference paper; |
| Johnson 1996 | Characterization of a new Marburg virus isolated from a 1987 fatal case in Kenya | Archives of virology. Supplementum | Exclusion reason: Not peer-reviewed paper; JS (2019-08-09 18:41:51)(Select): exclude, not a peer-reviewed paper; JS (2019-06-28 00:28:18)(Select): conference paper; JS (2019-03-19 20:19:33)(Screen): case study; |
| Johnson 1982 | Marburg, Ebola and Rift Valley Fever virus antibodies in East African primates | Transactions of the Royal Society of Tropical Medicine and Hygiene | Exclusion reason: Wrong pathogen or pathogen epidemiology or transmission not main focus; JS (2019-08-23 20:56:56)(Select): Has seroprevalence amongst human animal handlers; JS (2019-07-30 00:38:12)(Select): animal handlers were all Marburg -; JS (2019-03-19 20:10:14)(Screen): seroprevalence in animals; |
| Jeffs 2007 | The Medecins Sans Frontieres intervention in the Marburg hemorrhagic fever epidemic, Uige, Angola, 2005. I. Lessons learned in the hospital | The Journal of infectious diseases | Exclusion reason: No report of parameters (including seroprevalence and other measures of interest) or transmission models or historical outbreaks; |

Table S8: Excluded studies at full text review with exclusion reason

| Study | Title | Journal | Notes |
|---|---|---|---|
| Jeffs 2006 | A clinical guide to viral haemorrhagic fevers: Ebola, Marburg and Lassa | Tropical doctor | Exclusion reason: Wrong pathogen or pathogen epidemiology or transmission not main focus; JS (2019-07-29 21:12:02)(Select): Deals with Ebola and Marburg together, gives both same incubation period etc. Not really Marburg-specific. Marking as wrong pathogen; JS (2019-03-19 20:08:52)(Screen): may have info on CFR, incubation period etc.; |
| "Isaacson 2001 | Viral hemorrhagic fever hazards for travelers in Africa | Clinical infectious diseases : an official publication of the Infectious Diseases Society of America | Exclusion reason: Wrong pathogen or pathogen epidemiology or transmission not main focus; JS (2019-07-29 21:00:17)(Select): Deals with Marburg and Ebola together, not very helpful. Tagged as wrong pathogen etc. because of the joining of the two ; |
| " Colebunders 2007 | Marburg hemorrhagic fever in Durba and Watsa, Democratic Republic of the Congo: clinical documentation, features of illness, and treatment | The Journal of infectious diseases | Exclusion reason: Reports metrics from other papers (not original estimates of primary data); JS (2019-07-18 19:03:52)(Select): CFR split by whether or not patients received healthcare; JS (2019-07-18 19:02:36)(Select): Onset to symptoms; |
| Colebunders 2004 | Organisation of health care during an outbreak of Marburg haemorrhagic fever in the Democratic Republic of Congo, 1999 | The Journal of infection | Exclusion reason: No report of parameters (including seroprevalence and other measures of interest) or transmission models or historical outbreaks; JS (2019-07-18 18:51:33)(Select): The outbreak in this paper is at least partially covered by other papers (eg. the one with the infant by Borchert et al.). I think that this paper does not really have new info about the transmission of that outbreak and they do not provide risk factors. I'll include for now just in case we don't have all the references in this paper in the review list. ; |
| Changula 2014 | Ebola and Marburg virus diseases in Africa: increased risk of outbreaks in previously unaffected areas? | Microbiology and immunology | Exclusion reason: Reports metrics from other papers (not original estimates of primary data); JS (2019-03-15 21:07:35)(Screen): Mention of outbreaks; |
| Callendret 2018 | A prophylactic multivalent vaccine against different filovirus species is immunogenic and provides protection from lethal infections with Ebolavirus and Marburgvirus species in non-human primates | PloS one | Exclusion reason: No report of parameters (including seroprevalence and other measures of interest) or transmission models or historical outbreaks; JS (2019-03-19 00:53:30)(Screen): CFR in primates?; |
| Burton 2004 | Marburg miner mystery | Lancet Infect Dis | Exclusion reason: Not peer-reviewed paper; JS (2019-06-27 22:16:07)(Select): news; |
| Brown 1997 | Threat to Humans from Virus Infections of Non-human Primates | Reviews in medical virology | Exclusion reason: Wrong pathogen or pathogen epidemiology or transmission not main focus; JS (2019-07-18 00:09:51)(Select): Not worth keeping - everything here can be found elsewhere and Marburg not the main focus; JS (2019-03-15 20:53:38)(Screen): Mention of outbreak; |
| Brett-Major 2018 | Catching Chances: The Movement to Be on the Ground and Research Ready before an Outbreak | Viruses | Exclusion reason: Wrong pathogen or pathogen epidemiology or transmission not main focus; |
| Brauburger 2012 | Forty-five years of Marburg virus research | Viruses | Exclusion reason: Reports metrics from other papers (not original estimates of primary data); |

Table S8: Excluded studies at full text review with exclusion reason

| Study | Title | Journal | Notes |
|---|---|---|---|
| "Bramble 2018 | Pan-Filovirus Serum Neutralizing Antibodies in a Subset of Congolese Ebolavirus Infection Survivors | The Journal of infectious diseases | Exclusion reason: Wrong pathogen or pathogen epidemiology or transmission not main focus; JS (2019-07-17 23:49:42)(Select): This paper is about Ebola survivors, not Marburg infection ; |
| " Brainard 2016 | Presence and Persistence of Ebola or Marburg Virus in Patients and Survivors: A Rapid Systematic Review | PLoS neglected tropical diseases | Exclusion reason: Wrong pathogen or pathogen epidemiology or transmission not main focus; JS (2019-07-17 23:47:32)(Select): Mostly about Ebola; JS (2019-03-15 20:21:18)(Screen): Review may contain useful references; |
| Brainard 2016 | Risk factors for transmission of Ebola or Marburg virus disease: a systematic review and meta-analysis | International journal of epidemiology | Exclusion reason: Reports metrics from other papers (not original estimates of primary data); JS (2019-07-17 21:27:14)(Select): Risk factors possibly in the relevant references; |
| Borchert 2005 | Lessons from the outbreak of Marburg virus | N Engl J Med | Exclusion reason: Not peer-reviewed paper; |
| Borchert 2000 | Viewpoint: filovirus haemorrhagic fever outbreaks: much ado about nothing? | Tropical medicine | international health : TM |
| IH | Exclusion reason: No report of parameters (including seroprevalence and other measures of interest) or transmission models or historical outbreaks; | | |
| Bonney 2013 | Hospital-based surveillance for viral hemorrhagic fevers and hepatitides in Ghana | PLoS neglected tropical diseases | Exclusion reason: No report of parameters (including seroprevalence and other measures of interest) or transmission models or historical outbreaks; CM (2023-05-01 02:28:54)(Select): We have other sero-studies which report 0 prevalence ...; JS (2019-03-15 00:59:52)(Screen): Found a seroprevalence of 0 in humans; |
| "Bonn 2005 | Marburg fever in Angola: still a mystery disease | Lancet Infect Dis | Exclusion reason: Not peer-reviewed paper; JS (2019-03-15 00:34:02)(Screen): Not really a journal article - more of an interview ; |
| " Beer 1999 | Characteristics of Filoviridae: Marburg and Ebola viruses | Die Naturwissenschaften | Exclusion reason: Reports metrics from other papers (not original estimates of primary data); JS (2019-03-14 23:15:16)(Screen): Info on Marburg outbreaks and case fatality; |
| Bebell 2015 | Ebola virus disease and Marburg disease in pregnancy: a review and management considerations for filovirus infection | Obstetrics and gynecology | Exclusion reason: Reports metrics from other papers (not original estimates of primary data); JS (2019-03-14 23:08:12)(Screen): Review; |
| Bausch 2008 | Treatment of Marburg and Ebola hemorrhagic fevers: a strategy for testing new drugs and vaccines under outbreak conditions | Antiviral research | Exclusion reason: Reports metrics from other papers (not original estimates of primary data); JS (2019-03-15 00:05:01)(Screen): Case fatality rate mentioned - may have a reference for it; |
| Barrette 2011 | Current perspectives on the phylogeny of Filoviridae | Infection, genetics and evolution : journal of molecular epidemiology and evolutionary genetics in infectious diseases | Exclusion reason: No report of parameters (including seroprevalence and other measures of interest) or transmission models or historical outbreaks; JS (2019-03-15 00:09:59)(Screen): Mentions outbreaks and Marburg - may have relevant references; |

Table S8: Excluded studies at full text review with exclusion reason

| Study | Title | Journal | Notes |
|---|---|---|---|
| "Bannister 2010 | Viral haemorrhagic fevers imported into non-endemic countries: risk assessment and management | British medical bulletin | Exclusion reason: Wrong pathogen or pathogen epidemiology or transmission not main focus; JS (2019-03-14 23:05:01)(Screen): Not solely about filoviruses ; |
| " Ascenzi 2008 | Ebolavirus and Marburgvirus: insight the Filoviridae family | Molecular aspects of medicine | Exclusion reason: Reports metrics from other papers (not original estimates of primary data); JS (2019-03-14 22:27:02)(Screen): Looks like a review - may have useful references; |
| Amman 2014 | Marburgvirus resurgence in Kitaka Mine bat population after extermination attempts, Uganda | Emerging infectious diseases | Exclusion reason: No report of parameters (including seroprevalence and other measures of interest) or transmission models or historical outbreaks; |
| Amman 2015 | Oral shedding of Marburg virus in experimentally infected Egyptian fruit bats (Rousettus aegyptiacus) | Journal of wildlife diseases | Exclusion reason: No report of parameters (including seroprevalence and other measures of interest) or transmission models or historical outbreaks; JS (2019-03-14 22:18:34)(Screen): May contain human outbreak info; |
| "Alves 2010 | Aerosol exposure to the angola strain of marburg virus causes lethal viral hemorrhagic Fever in cynomolgus macaques | Veterinary pathology | Exclusion reason: Wrong pathogen or pathogen epidemiology or transmission not main focus; JS (2019-03-14 21:55:37)(Screen): may get an animal estimate of incubation period ; |
| " Allaranga 2010 | Lessons learned during active epidemiological surveillance of Ebola and Marburg viral hemorrhagic fever epidemics in Africa | East African journal of public health | Exclusion reason: Reports metrics from other papers (not original estimates of primary data); JS (2019-06-17 19:27:59)(Select): Contains list of Marburg outbreaks - though no original data. Good to retain for reference-checking?; |
| Alfson 2018 | A Single Amino Acid Change in the Marburg Virus Glycoprotein Arises during Serial Cell Culture Passages and Attenuates the Virus in a Macaque Model of Disease | mSphere | Exclusion reason: No report of parameters (including seroprevalence and other measures of interest) or transmission models or historical outbreaks; JS (2019-03-13 04:05:06)(Screen): Can possibly calculate CFR in animal model from study data; |
| Adjemian 2011 | Outbreak of Marburg hemorrhagic fever among miners in Kamwenge and Ibanda Districts, Uganda, 2007 | The Journal of infectious diseases | Exclusion reason: Duplicate; JS (2019-03-13 03:48:12)(Screen): Duplicate; JS (2019-03-13 03:45:48)(Screen): Outbreak; |
| | Case definitions. Ebola-Marburg viral diseases | Epidemiological bulletin | Exclusion reason: Not peer-reviewed paper; |
| | After Marburg, Ebola | Lancet | Exclusion reason: Wrong pathogen or pathogen epidemiology or transmission not main focus; JS (2019-08-24 01:50:42)(Select): About Ebola; |
| Polonsky 2014 | Perspective Piece Emerging Filoviral Disease in Uganda: Proposed Explanations and Research Directions | American Journal of Tropical Medicine and Hygiene | Exclusion reason: Duplicate; JS (2019-03-20 19:42:35)(Screen): duplicate; |
| Okeke 2014 | Diagnostic schemes for reducing epidemic size of african viral hemorrhagic fever outbreaks | Journal of Infection in Developing Countries | Exclusion reason: Wrong pathogen or pathogen epidemiology or transmission not main focus; JS (2019-08-01 00:36:24)(Select): The model seems to be parameterised and used on only Ebola; JS (2019-03-20 02:44:44)(Screen): Focuses on Ebola but paper may also consider Marburg?; |

Table S8: Excluded studies at full text review with exclusion reason

| Study | Title | Journal | Notes |
|---|---|---|---|
| Nyakarahuka 2014 | Using network analysis technique to describe the spread of Marburg hemorrhagic fever outbreak in Uganda, 2012 | International Journal of Infectious Diseases | Exclusion reason: Not peer-reviewed paper; JS (2019-07-09 20:50:31)(Select): Oral presentation; |
| Nakayama 2011 | Ebola and Marburg viruses | Journal of Disaster Research | Exclusion reason: Reports metrics from other papers (not original estimates of primary data); JS (2019-03-19 23:17:23)(Screen): mention of outbreaks; |
| Kupradze 1981 | VIRAL HEMORRHAGIC FEVERS | Izvestiya Akademii Nauk Gruzinskoi SSR Seriya Biologicheskaya | Exclusion reason: Not in English; |
| Kortepeter 2011 | Basic Clinical and Laboratory Features of Filoviral Hemorrhagic Fever | Journal of Infectious Diseases | Exclusion reason: Duplicate; JS (2019-03-19 20:56:53)(Screen): duplicate; |
| Rodhain 1989 | ARBOVIRUS INFECTIONS AND VIRAL HEMORRHAGIC FEVERS IN UGANDA - A SEROLOGICAL SURVEY IN KARAMOJA DISTRICT, 1984 | Transactions of the Royal Society of Tropical Medicine and Hygiene | Exclusion reason: Duplicate; JS (2019-03-15 01:02:03)(Screen): Strictly speaking not Marburg alone, but useful for serological data; |
| Roddy 2007 | The medecins sans frontieres intervention in the Marburg hemorrhagic fever epidemic, Uige, Angola, 2005. II. lessons learned in the community | Journal of Infectious Diseases | Exclusion reason: Duplicate; JS (2019-03-20 19:58:19)(Screen): duplicate; |
| Roddy 2009 | Decreased peripheral health service utilisation during an outbreak of Marburg haemorrhagic fever, Uige, Angola, 2005 | Transactions of the Royal Society of Tropical Medicine and Hygiene | Exclusion reason: Reports metrics from other papers (not original estimates of primary data); JS (2019-08-05 23:35:41)(Select): Not really about Marburg in itself; |
| Knust 2015 | Multidistrict Outbreak of Marburg Virus Disease-Uganda, 2012 | Journal of Infectious Diseases | Exclusion reason: Duplicate; JS (2019-03-19 20:51:53)(Screen): duplicate; |
| "Kalter 1971 | A SEROLOGICAL SURVEY OF PRIMATE SERA FOR ANTIBODY TO THE MARBURG VIRUS | Martini, G. a. and R. Siegert | Exclusion reason: Not peer-reviewed paper; JS (2019-07-09 18:08:11)(Select): Book chapter; JS (2019-03-13 04:08:23)(Screen): Also contains seroprevalence data in humans - humans are primates ; |
| " Johnson 1996 | Characterization of a new Marburg virus isolated from a 1987 fatal case in Kenya | Archives of Virology Supplement | Exclusion reason: Duplicate; JS (2019-03-19 20:19:15)(Screen): duplicate; |
| Johnson 1993 | HEMORRHAGIC-FEVER VIRUS ACTIVITY IN EQUATORIAL AFRICA - DISTRIBUTION AND PREVALENCE OF FILOVIRUS REACTIVE ANTIBODY IN THE CENTRAL-AFRICAN-REPUBLIC | Transactions of the Royal Society of Tropical Medicine and Hygiene | Exclusion reason: Duplicate; JS (2019-03-19 20:11:34)(Screen): seroprevalence in humans; |

Table S8: Excluded studies at full text review with exclusion reason

| Study | Title | Journal | Notes |
|---|---|---|---|
| Johnson 1983 | VIRAL HEMORRHAGIC-FEVER SURVEILLANCE IN KENYA, 1980-1981 | Tropical and Geographical Medicine | Exclusion reason: Duplicate; JS (2019-03-19 20:10:51)(Screen): duplicate; |
| Jeffs 2007 | The medecins sans frontieres intervention in the Marburg hemorrhagic fever epidemic, Uige, Angola, 2005. I. lessons learned in the hospital | Journal of Infectious Diseases | Exclusion reason: Duplicate; JS (2019-03-19 20:09:17)(Screen): duplicate; |
| Ivanoff 1982 | HEMORRHAGIC-FEVER IN GABON .1. INCIDENCE OF LASSA, EBOLA AND MARBURG VIRUSES IN HAUT-OGOOUE | Transactions of the Royal Society of Tropical Medicine and Hygiene | Exclusion reason: Duplicate; JS (2019-03-19 03:59:55)(Screen): seroprevalence?; |
| Gonzalez 1983 | AFRICAN VIRAL HEMORRHAGIC FEVERS STUDIES IN THE CENTRAL-AFRICAN-REPUBLIC | Cahiers O.R.S.T.O.M. (Office de la Recherche Scientifique et Technique Outre-Mer) Serie Entomologie Medicale et Parasitologie | Exclusion reason: Not in English; |
| Towner 2007 | High-throughput molecular detection of hemorrhagic fever virus threats with applications for outbreak settings | Journal of Infectious Diseases | Exclusion reason: No report of parameters (including seroprevalence and other measures of interest) or transmission models or historical outbreaks; JS (2019-08-08 23:02:40)(Select): Did analysis of 500 people for Uige outbreak – 180+/505; JS (2019-03-21 20:23:53)(Screen): may contain data on original seroprevalence in these 500 people; |
| Sureau 1989 | Recent findings on the African viral haemorrhagic fevers | Maladies tropicales transmissibles. | Exclusion reason: Not in English; JS (2019-03-20 21:44:43)(Screen): may be in French?; |
| Slenczka 2007 | Forty years of Marburg virus | Journal of Infectious Diseases | Exclusion reason: Reports metrics from other papers (not original estimates of primary data); |
| Martini 1971 | MARBURG VIRUS DISEASE CLINICAL SYNDROME | Martini, G. a. and R. Siegert | Exclusion reason: Not peer-reviewed paper; JS (2019-07-09 18:35:24)(Select): Book chapter; KC (2019-03-19 22:44:35)(Screen): incubation period from first outbreak reported here; |
| Lub 1995 | Clinical and virologic characterization of the disease in guinea pigs aerogenically infected with Marburg virus | Voprosy Virusologii | Exclusion reason: Not in English; JS (2019-03-19 21:46:23)(Screen): incubation period etc. in guinea pigs; |
| Ftika 2013 | Viral haemorrhagic fevers in healthcare settings | Journal of Hospital Infection | Exclusion reason: Wrong pathogen or pathogen epidemiology or transmission not main focus; JS (2019-07-19 23:32:31)(Select): Review talks a lot about Ebola and Marburg together. Not really Marburg-specific; |
| Freitas 2017 | Filovirus strains, the environment conditions and the bats in and out of Africa | Archives of Veterinary Science | Exclusion reason: No report of parameters (including seroprevalence and other measures of interest) or transmission models or historical outbreaks; JS (2019-03-18 23:14:01)(Screen): outbreak range; |
| Formenty 2006 | Viral haemorrhagic fevers in the world: review of the last ten years | Bulletin Epidemiologique Hebdomadaire | Exclusion reason: Not in English; JS (2019-10-18 02:13:23)(Select): Requested from library 17/10/2019; |
| Ford 1999 | Haemorrhagic fever in Democratic Republic of Congo identified as Marburg | Lancet | Exclusion reason: Not peer-reviewed paper; JS (2019-06-27 23:00:07)(Select): news; |

Table S8: Excluded studies at full text review with exclusion reason

| Study | Title | Journal | Notes |
|---|---|---|---|
| Fernando 2015 | Immune Response to Marburg Virus Angola Infection in Non-human Primates | Journal of Infectious Diseases | Exclusion reason: Duplicate; |
| Feldmann 2006 | Focus on research: Marburg hemorrhagic fever - The forgotten cousin strikes | New England Journal of Medicine | Exclusion reason: Reports metrics from other papers (not original estimates of primary data); JS (2019-07-18 19:49:59)(Select): Perspective – I think we're excluding these; |
| Feldmann 1996 | Emerging and reemerging of filoviruses | Archives of Virology | Exclusion reason: Not peer-reviewed paper; JS (2019-07-11 19:47:59)(Select): Book chapter; |
| Farnon 2009 | FILOVIRUS SEROURVEY FOLLOWING AN OUTBREAK OF MARBURG HEMORRHAGIC FEVER — IBANDA AND KAMWENGE DISTRICTS, UGANDA, 2007 | American Journal of Tropical Medicine and Hygiene | Exclusion reason: Not peer-reviewed paper; JS (2019-08-09 18:40:33)(Select): conference proceedings - exclude; |
| Falzarano 2006 | Characterization of Marburg virus from a recent outbreak in Angola | American Journal of Tropical Medicine and Hygiene | Exclusion reason: Not peer-reviewed paper; JS (2019-08-09 18:40:17)(Select): conference proceedings - exclude; |
| Eichenlaub 1985 | HEMORRHAGIC FEVERS RISK FOR HOSPITAL STAFF | Hygiene + Medizin | Exclusion reason: Not in English; |
| Dietrich 1978 | Marburg virus disease | Ebola virus haemorrhagic fever. Proceedings of an international colloquium on Ebola virus infection and other haemorrhagic fevers held in Antwerp, Belgium, 6-8 December, 1977. | Exclusion reason: Not peer-reviewed paper; JS (2019-06-27 22:34:53)(Select): Proceedings of an International Colloquium; |
| Colebunders 2007 | Marburg hemorrhagic fever in durba and watsa, democratic republic of the congo: Clinical documentation, features of illness, and treatment | Journal of Infectious Diseases | Exclusion reason: Duplicate; |
| Tessier 1987 | VIRAL HEMORRHAGIC-FEVER SURVEY IN CHOBE (BOTSWANA) | Transactions of the Royal Society of Tropical Medicine and Hygiene | Exclusion reason: Wrong pathogen or pathogen epidemiology or transmission not main focus; JS (2019-08-08 20:00:16)(Select): zero prevalence of Marburg; |
| Hibbs 1993 | Epidemic of febrile disease in Berbera, Somalia | Journal of Tropical Medicine | Exclusion reason: No report of parameters (including seroprevalence and other measures of interest) or transmission models or historical outbreaks; JS (2019-03-19 03:38:52)(Screen): seroprevalence in humans?; |
| Hennessen 1971 | EPIDEMIOLOGY OF MARBURG VIRUS DISEASE | Martini, G. a. and R. Siegert | Exclusion reason: Not peer-reviewed paper; JS (2019-07-29 20:01:17)(Select): Book chapter?; |
| Henderson 1971 | EPIDEMIOLOGICAL STUDIES IN UGANDA RELATING TO THE MARBURG AGENT | Martini, G. a. and R. Siegert | Exclusion reason: Not peer-reviewed paper; JS (2019-07-29 20:01:03)(Select): Book chapter?; |
| Chepurnova 2000 | Assay of Marburg virus in the blood and secretions of experimentally infected animals | Voprosy Virusologii | Exclusion reason: Not in English; JS (2019-03-15 00:06:20)(Screen): Animal incubation period; |

Table S8: Excluded studies at full text review with exclusion reason

| Study | Title | Journal | Notes |
|---|---|---|---|
| Cdc 2005 | Brief report: Outbreak of Marburg virus hemorrhagic fever - Angola, October 1, 2004-March 29, 2005 (Reprinted from MMWR, vol 54, pg 308-309, 2005) | Jama-Journal of the American Medical Association | Exclusion reason: Not peer-reviewed paper; |
| Cardenas 2013 | Marburg marburgvirus | Mononegaviruses of veterinary importance. Volume I: Pathobiology and molecular diagnosis | Exclusion reason: Not peer-reviewed paper; JS (2019-06-27 22:18:25)(Select): book chapter; |
| Buchmeier 1984 | MARBURG AND EBOLA VIRUSES NEW AGENTS ON THE FRONTIERS OF VIROLOGY | Notkins, a. L. and M. B. a. Oldstone | Exclusion reason: Not peer-reviewed paper; JS (2019-06-27 22:19:14)(Select): book chapter; JS (2019-03-15 20:57:05)(Screen): Mention of historical outbreaks; |
| Belanov 1996 | Survival of Marburg virus on contaminated surfaces and in aerosol | Voprosy Virusologii | Exclusion reason: Not in English; JS (2019-05-17 19:59:07)(Select): In Russian; |
| Bausch 1999 | Investigation of an outbreak of Marburg hemorrhagic fever in the Democratic Republic of Congo | American Journal of Tropical Medicine and Hygiene | Exclusion reason: Not peer-reviewed paper; JS (2019-08-08 23:57:12)(Select): Conference proceedings - not a journal article; JS (2019-05-17 19:53:23)(Select): Could not find the paper; |
| Anonymous 2005 | Outbreak of Marburg virus hemorrhagic fever - Angola, October 1, 2004-March 29, 2005 | Morbidity and Mortality Weekly Report | Exclusion reason: Case report or case study (i.e. reports on less than 10 cases, but this threshold can be pathogen-dependent); JS (2019-06-26 23:54:47)(Select): This report is from the middle of the outbreak, so technically does not report parameters of interest; |
| Amman 2017 | Ecology of Filoviruses | Marburg- and Ebolaviruses: from Ecosystems to Molecules | Exclusion reason: Not peer-reviewed paper; JS (2019-05-17 19:44:39)(Select): Book chapter; |
| Amdiouni 2015 | Ebola virus and other Filoviruses: an overview | Journal of Coastal Life Medicine | Exclusion reason: Reports metrics from other papers (not original estimates of primary data); KC (2019-07-02 18:47:26)(Select): mostly Ebola; JS (2019-03-14 21:57:29)(Screen): About filoviruses - may contain a review of Marburg?; |
| Borchert 2002 | A cluster of Marburg virus disease involving an infant | Tropical Medicine | International Health |
| Exclusion reason: Duplicate; | | | |
| Boardman 2003 | Viral hemorrhagic fever | Primary Care Update for Ob/Gyns | Exclusion reason: Reports metrics from other papers (not original estimates of primary data); JS (2019-03-15 00:29:36)(Screen): incubation period estimate; |
| "Adegboro 2011 | Marburg haemorrhagic fever: recent advances | African Journal of Clinical and Experimental Microbiology | Exclusion reason: No report of parameters (including seroprevalence and other measures of interest) or transmission models or historical outbreaks; JS (2019-03-13 03:40:08)(Screen): A review? ; |
| " | Outbreak news. Marburg haemorrhagic fever, Uganda | Releve epidemiologique hebdomadaire | Exclusion reason: Not peer-reviewed paper; JS (2019-03-20 23:58:32)(Screen): Not really a journal article but adding it in anyways because it's kind of a case; |
| | Largest marburg outbreak ever recorded hits Angola | Biosecurity and Bioterrorism-Biodefense Strategy Practice and Science | Exclusion reason: Not peer-reviewed paper; JS (2019-08-08 23:55:45)(Select): This seems to not really be a journal article, but a bunch of summaries? Probably not a peer-reviewed journal article?; |
| Adepoju 2021 | West Africa on alert for haemorrhagic fevers | Lancet | Exclusion reason: Case report or case study (i.e. reports on less than 10 cases, but this threshold can be pathogen-dependent); |

Table S8: Excluded studies at full text review with exclusion reason

| Study | Title | Journal | Notes |
|---|---|---|---|
| Araf 2023 | Marburg virus outbreak in 2022: a public health concern | Lancet Microbe | Exclusion reason: No report of parameters (including seroprevalence and other measures of interest) or transmission models or historical outbreaks; |
| Asad 2020 | Past and current advances in Marburg virus disease: a review | Infez Med | Exclusion reason: Reports metrics from other papers (not original estimates of primary data); |
| Baby 2022 | Sagacious perceptive on Marburg virus foregrounding the recent findings :A Critical Review | Infect Disord Drug Targets | Exclusion reason: Reports metrics from other papers (not original estimates of primary data); |
| Bouba 2023 | Predicting the combined effects of case isolation, safe funeral practices, and contact tracing during Ebola virus disease outbreaks | PLoS One | Exclusion reason: Wrong pathogen or pathogen epidemiology or transmission not main focus; |
| Chakraborty 2022 | Sexual transmission of recently re-emerged deadly Marburg virus (MARV) needs explorative studies and due attention for its prevention and feasible spread - Correspondence | Int J Surg | Exclusion reason: No report of parameters (including seroprevalence and other measures of interest) or transmission models or historical outbreaks; |
| Filion 2023 | Preliminary Investigation of Schmalhausen's Law in a Directly Transmitted Pathogen Outbreak System | Viruses | Exclusion reason: No report of parameters (including seroprevalence and other measures of interest) or transmission models or historical outbreaks; |
| Fujita-Fujiharu 2022 | Structural insight into Marburg virus nucleoprotein-RNA complex formation | Nat Commun | Exclusion reason: No report of parameters (including seroprevalence and other measures of interest) or transmission models or historical outbreaks; |
| Harris 2023 | WHO: Marburg Virus Outbreak Confirmed in Equatorial Guinea | Jama | Exclusion reason: Not peer-reviewed paper; CM (2023-05-01 19:43:17)(Select): Case report for Equatorial Guinea. Should we include?; |
| Hunter 2023 | Marburg Fever | StatPearls | Exclusion reason: No report of parameters (including seroprevalence and other measures of interest) or transmission models or historical outbreaks; |
| Iannetta 2019 | Viral Hemorrhagic Fevers Other than Ebola and Lassa | Infect Dis Clin North Am | Exclusion reason: Reports metrics from other papers (not original estimates of primary data); |
| Islam 2023 | Epidemiology, pathophysiology, transmission, genomic structure, treatment, and future perspectives of the novel Marburg virus outbreak | Int J Surg | Exclusion reason: Case report or case study (i.e. reports on less than 10 cases, but this threshold can be pathogen-dependent); |
| Jacobs 2023 | They come in threes: Marburg virus, emerging infectious diseases, and the blood supply | Transfus Apher Sci | Exclusion reason: Not peer-reviewed paper; CM (2023-05-01 19:38:43)(Select): This would only be recording the (recent) outbreak with 2 cases in Ghana in 2022; |
| Janik 2020 | Dangerous Pathogens as a Potential Problem for Public Health | Medicina (Kaunas) | Exclusion reason: Wrong pathogen or pathogen epidemiology or transmission not main focus; |

Table S8: Excluded studies at full text review with exclusion reason

| Study | Title | Journal | Notes |
|---|---|---|---|
| Jonkmans 2021 | Scoping future outbreaks: a scoping review on the outbreak prediction of the WHO Blueprint list of priority diseases | BMJ Glob Health | Exclusion reason: No report of parameters (including seroprevalence and other measures of interest) or transmission models or historical outbreaks; |
| Koundouno 2022 | Detection of Marburg Virus Disease in Guinea | N Engl J Med | Exclusion reason: Case report or case study (i.e. reports on less than 10 cases, but this threshold can be pathogen-dependent); |
| Languon 2019 | Filovirus Disease Outbreaks: A Chronological Overview | Virology (Auckl) | Exclusion reason: No report of parameters (including seroprevalence and other measures of interest) or transmission models or historical outbreaks; |
| Mohapatra 2022 | Recent re-emergence of Marburg virus disease in an African country Ghana after Guinea amid the ongoing COVID-19 pandemic: Another global threat? Current knowledge and strategies to tackle this highly deadly disease having feasible pandemic potential | Int J Surg | Exclusion reason: No report of parameters (including seroprevalence and other measures of interest) or transmission models or historical outbreaks; |
| Olejnik 2019 | Recent advances in marburgvirus research | F1000Res | Exclusion reason: Reports metrics from other papers (not original estimates of primary data); |
| Ristanovifá 2020 | A Forgotten Episode of Marburg Virus Disease: Belgrade, Yugoslavia, 1967 | Microbiol Mol Biol Rev | Exclusion reason: Reports metrics from other papers (not original estimates of primary data); CM (2023-05-01 02:32:02)(Select): Includes parameter estimates from prior non-english studies; |
| Sah 2022 | Marburg virus re-emerged in 2022: recently detected in Ghana, another zoonotic pathogen coming up amid rising cases of Monkeypox and ongoing COVID-19 pandemic- global health concerns and counteracting measures | Vet Q | Exclusion reason: No report of parameters (including seroprevalence and other measures of interest) or transmission models or historical outbreaks; |
| Sahoo 2023 | The Marburg virus outbreak in West Africa | Curr Drug Targets | Exclusion reason: Not peer-reviewed paper; |
| Shifflett 2019 | Marburg virus pathogenesis - differences and similarities in humans and animal models | Virol J | Exclusion reason: Reports metrics from other papers (not original estimates of primary data); |
| Stephens 2022 | Drivers of African Filovirus (Ebola and Marburg) Outbreaks | Vector Borne Zoonotic Dis | Exclusion reason: No report of parameters (including seroprevalence and other measures of interest) or transmission models or historical outbreaks; |
| Tahmo 2023 | An epidemiological synthesis of emerging and re-emerging zoonotic disease threats in Cameroon, 2000-2022: a systematic review | IJID Reg | Exclusion reason: No report of parameters (including seroprevalence and other measures of interest) or transmission models or historical outbreaks; |
| Woolsey 2020 | Immune correlates of postexposure vaccine protection against Marburg virus | Sci Rep | Exclusion reason: No report of parameters (including seroprevalence and other measures of interest) or transmission models or historical outbreaks; |

Table S8: Excluded studies at full text review with exclusion reason

| Study | Title | Journal | Notes |
|---|---|---|---|
| Mashkoor 2022 | Recurrent Marburg virus disease outbreaks from 1967 to 2022: A perspective on challenges imposed and future implications | ASIAN PACIFIC JOURNAL OF TROPICAL MEDICINE | Exclusion reason: No report of parameters (including seroprevalence and other measures of interest) or transmission models or historical outbreaks; |

Table S8: Excluded studies at full text review with exclusion reason
